# Comparison of Single Polygenic, Multiple Polygenic Risk, and Lifestyle for Brain Health Index in Explaining Cognitive Function Among Middle-aged and Older Adults in The Maastricht Study

**DOI:** 10.1101/2025.06.16.25329671

**Authors:** Yidan Zhang, Sinan Gülöksüz, Coen Stehouwer, Marleen van Greevenbroek, Sebastian Köhler, Simone J.P.M. Eussen, Hans Bosma, Martin P.J. van Boxtel, Miranda Schram, David E.J. Linden, Bart P.F. Rutten, Gabriëlla Blokland

## Abstract

Cognitive function is shaped by both genetic and environmental factors. The Lifestyle for Brain Health (LIBRA) index, based on epidemiological evidence, targets modifiable risk and protective factors during midlife and early old age. This study compares the explanatory power of polygenic risk scores (PGSs) and the LIBRA score in relation to cognitive function among middle-aged and older adults in the Maastricht Study.

We analyzed 17 cognition-related PGSs individually and combined significant PGSs into a multi-PGS model. The performance of the LIBRA model, individual PGS models, the multi-PGS model, and integrated LIBRA-genotype models was evaluated. The intelligence PGS exhibited the strongest association with cognitive function (β = 0.109, 95% CI: 0.094-0.124). Five PGSs remained significant and were incorporated into the multi-PGS model. Compared to the LIBRA-only model, genetic models, including either the top single-PGS or multi-PGS, showed improved performance, with Adjusted R² increasing by 2.5% to 3.1%. The LIBRA + multi-PGS model provided the highest explanatory power, with a 4% increase in Adjusted R², validated by 10-fold cross-validation.

These results underscore the value of integrating PGSs, particularly multi-PGS models, with the LIBRA score to enhance the prediction of cognitive outcomes. This genetic-environmental approach offers potential for better understanding and predicting cognitive function in middle-aged to early old-aged populations, with implications for clinical and public health applications.

## Introduction

Cognition is a multifaceted construct, encompassing domains such as memory, information processing speed, and executive functions; all cognitive processes that contribute to individual differences in intelligence [1]. Cognitive abilities are essential for daily functioning and quality of life. The increasing number of adults over 65 and the rising prevalence of age-associated neurodegenerative dementias underscore the growing importance of cognitive health in our aging society [2]. Therefore, exploring factors that predict cognitive changes and identifying effective preventative or therapeutic strategies to preserve cognitive function in advanced age are of paramount importance.

Recent advances in neuroscience and epidemiology have highlighted both genetic predispositions and lifestyle factors as key determinants of cognitive health [3]. Studies focusing on genetic predispositions to cognitive functions have revealed heritability estimates ranging from 40-80% [4,5,6,7]. Simultaneously, studies have identified numerous modifiable risk factors associated with cognitive health, encompassing lifestyle choices, mental health conditions, and chronic diseases. These findings suggest that interventions targeting such factors could play a critical role in both mitigating cognitive decline and enhancing cognitive function, particularly in middle-aged and older adults [8,9,10]. To provide a clinical benchmark for cognitive risk assessment, researchers have developed tools like the LIfestyle for BRAin health (LIBRA) score which, based on a systematic literature review and Delphi consensus, provides a weighted composite score to quantitatively assess modifiable risk factors associated with cognitive decline [8]. Beyond its correlation with cognitive decline, the LIBRA score has been validated and shown to possess significant explanatory value for cognitive functions, especially in the context of aging [11,12,13]. Notably, each one-point increment in the LIBRA score corresponds to a 19% increase in the risk for dementia and a 9% increase in the risk for cognitive impairment [14]. These insights affirm the practical utility of the LIBRA score in pinpointing and monitoring an individual’s risk profile, emphasizing the pivotal role of modifiable, lifestyle-related risk factors not just in preventing cognitive impairment, but also in supporting cognitive function among middle-aged and older individuals.

Furthermore, recent advancements in genome-wide association studies (GWASs) have deepened our understanding of the genetic factors influencing cognitive function. In particular, large-scale GWASs have identified over 100 genome-wide significant loci related to cognitive function [15,16,17,18], which has facilitated the development of polygenic risk scores (PGSs) for identifying these genetic influences. These scores aggregate the weighted effects of numerous small genetic variations, offering a nuanced genetic risk assessment.

Cognitive functioning is interconnected with various traits such as mental health, neurodevelopmental disorders, cardiometabolic diseases, brain structure, and sleep patterns, all of which share certain genetic underpinnings with cognitive processes [16,19,20,21,22,23]. Studies have suggested that combining multiple PGSs relevant to the target trait enhances both explanatory power and predictive accuracy beyond what is achievable with individual PGS. This approach, known as the multi-PGS approach, has been shown to improve prediction precision and offer more comprehensive insights into the underlying biology of the trait under investigation [24,25].

Given these developments, our study seeks to explore how PGSs compare with the established LIBRA score, in their association with cognitive function among Middle-aged and Older people. We also investigate whether a combined approach, integrating genetic and LIBRA factors, can more accurately explain the proportion of cognitive outcomes. To achieve this objective, we constructed single PGS models by examining individual PGS associations with cognitive function and developed a comprehensive multi-PGS model by jointly incorporating multiple significant PGSs. Building on this foundation, we then evaluated how integrating PGSs (encompassing both single and multi-PGS models) with the LIBRA score enhances our ability to account for cognitive function, with the potential to enhance predictive accuracy and detection of cognitive function and impairment in Middle-aged and Older populations.

## Methods

### Participants

The current study utilizes data from The Maastricht Study (DMS), a longitudinal population-based study that focuses on the etiology, pathophysiology, complications, and comorbidities of type 2 diabetes (T2D). The rationale and methodology of the study have been previously described [26]. Participants aged 40 to 75 years who reside in the southern region of the Netherlands were recruited for the study through mass media campaigns, the use of municipal registries, and mailings from the regional Diabetes Patient Registry. In compliance with ethical standards, DMS has been approved by the institutional medical ethical committee (NL31329.068.10) and the Ministry of Health, Welfare, and Sports of the Netherlands (Permit 131088-105234-PG). Written informed consent was provided by all participants. DMS totally included cross-sectional data from 9,187 participants who completed baseline measurements between November 2010 and December 2017. In this study, we included 5,244 participants with genotyping data that passed quality control (QC) and had complete phenotypic information.

### Measures

#### Cognitive assessments

Cognitive function in DMS was assessed using a brief neuropsychological test battery [26]. Test scores were standardized and divided into three cognitive domains, i.e., memory function, executive function and attention, and information processing speed. Memory was evaluated using the Verbal Learning Test; information processing speed was assessed using the Stroop Color-Word Test Part I and II, the Concept Shifting Test Part A and B, and the Letter-Digit Substitution Test; executive function was evaluated using the Stroop Color-Word Test Part III and Concept Shifting Test Part C [27,28,29,30]; details of cognitive tests are in the **Supplementary Methods**. The overall cognitive function score was derived by taking the standardized average of the scores from the three domains. Individuals who scored ≥1.5 SDs below their norm-based expected score (age, gender, and education-matched norms) in any of the three cognitive domains were classified as having cognitive impairment [12]. In this study, our outcome measures encompassed overall cognition (primary), as well as the test scores of the three cognitive domains (secondary).

#### Genotyping and imputation

Genotyping was performed using the Illumina Infinium Global Screening Array BeadChip at Erasmus University Medical Center, Rotterdam, Netherlands, with a 95% initial success rate.Quality control and imputation were executed using the Rapid Imputation for COnsortia PipeLIne (RICOPILI) [31].Preliminary Quality control (QC) included checks for sex discrepancies, related samples, and strand-ambiguous SNPs, with further QC steps to ensure data accuracy and reliability, which all performed in Plink 1.9 [32]. Imputation was carried out using Eagle v2.3.5 [33] for prephasing and Minimac3 [34] with the 1000 Genomes Phase 3 reference panel [35]. Additional post-imputation QC steps were applied, including filtering for heterozygosity outliers, Hardy-Weinberg equilibrium, and minor allele frequency (MAF). Detailed QC procedures and imputation settings are described in the **Supplementary Methods.**

#### Polygenic risk scoring

PGSs were calculated using PRSice-2 [36] based on the publicly available summary statistics from GWASs of 17 different phenotypes, which were selected based on prior evidence for association of those phenotypes with cognitive function (see **Supplementary Table 2** for a full list of references). Seventeen PGSs were generated, namely for the phenotypes educational attainment (EA), intelligence quotient (IQ), Alzheimer’s disease (AD), major depressive disorder (MDD), anxiety disorder (ANX), schizophrenia (SCZ), bipolar disorder (BD), attention deficit/hyperactivity disorder (ADHD), autism spectrum disorder (ASD), T2D, coronary artery disease (CAD), brain surface area (SA), brain cortical thickness (CT), brain volume (BV), insomnia, sleep duration, and morningness (that is, being a morning person: yes/no). These PGSs were categorized into groups based on the traits they characterize, namely cognition, psychiatric disorders, neurodevelopmental disorders, cardiometabolic disease, brain structure and sleep.

PGSs were computed at evenly spaced p-value thresholds for the range of 5*10^−8^ − 0.5, to find the best-fitting PGS per GWAS dataset, i.e., the inclusion of SNPs in the PGS was chosen empirically. The best fitting PGS had the highest R^2^ value from linear regression, relating PGS to overall cognition. The explanatory power of the PGS derived from the GWAS was measured by the incremental *R^2^* statistic [37]. To account for the number of variables in the model, the incremental adjusted *R^2^* (*R^2^*adj) was reported primarily, which reflects the increase in the *R^2^*adj when the PGS is added to a regression model predicting the behavioral phenotype alongside a number of control variables (here: sex, age, genotyping batch, and 10 ancestry PCs). To enhance interpretability, all PGSs were standardized.

#### LIBRA index

The LIBRA index was employed to gauge the ability of modifiable risk factors to explain cognitive function. The factors for LIBRA were sourced from clinical data and were operationalized within DMS, except for one factor pertaining to ‘high cognitive activity’ (weight –3.2), which was unavailable [12]. The LIBRA total score was calculated by assigning weights to each factor based on relative risks obtained from published meta-analyses [38,39], with higher scores indicating a higher risk of dementia. Protective and risk factors encompassed in this study included adherence to dietary guidelines as measured by the Dutch Healthy Eating Index, low to moderate alcohol use, physical inactivity, smoking, obesity, depression, T2D, hypertension, hypercholesterolemia, heart disease, and chronic kidney disease. A comprehensive description of the LIBRA factors assigned weights, and operationalization in this dataset can be found in the **Supplementary Methods** and **Supplementary Table 3**.

### Statistical analyses

Descriptive statistics included mean (SD) and frequency (%); missing data were addressed using listwise deletion. Outliers in cognitive test scores, defined as exceeding four SDs from the mean, were treated via Winsorizing. Two-tailed tests with ⍺=0.05 were used. Pearson correlations assessed initial relationships among 17 PGSs.

To validate PGS robustness, each PGS was tested for significant association with its corresponding phenotype in DMS; 10 out of 17 PGSs had matching phenotypes. For PGS_EA_, educational level categories were used; PGS_IQ_ used overall cognition scores; PGS_AD_ used cognitive impairment status; PGS_MDD_ aligned with the Patient Health Questionnaire-9 (PHQ-9) scores [40]; PGS_ANX_ corresponded with anxiolytic medication use; PGS_ADHD_ used executive function and attention scores; PGS_T2D_ used T2D status; PGS_CAD_ used ‘history of cardiovascular disease’; PGS_insomnia_ used the sleep difficulty question; PGS_sleepduration_ used self-reported night sleep duration.

Cognition-related regression models were evaluated separately for Single PGS and Multi-PGS models. Single PGS models used linear regression with each PGS as an independent variable and cognitive variables (overall cognition, memory, processing speed, executive function) as dependent variables, adjusting for sex, age, genotyping batch, and 10 ancestry PCs. The Benjamini-Hochberg procedure [41] controlled the FDR across 17 PGSs. Significant PGSs corresponding to different cognitive variables were included in separate Multi-PGS models, each tailored to its respective dependent variable, to assess combined effects. Multicollinearity was checked via VIF [42] for each model.

The LIBRA-only model used regression analysis with total LIBRA score as the independent variable, and cognitive variables as dependent variables, adjusting for sex and age. We then integrated the most explanatory PGSs into the LIBRA model to test for performance improvement. Interaction terms for PGSs and LIBRA were also tested to assess their significance and potential inclusion.

Model performance was compared by evaluating overall explanatory power (R^2^ values) and the incremental R^2^ of key predictors (LIBRA and PGSs). A 10-fold cross-validation with 150 iterations using the ‘train’ function in the R package ‘caret’ was conducted, assessing three parameters: *R^2^*, Mean Absolute Error (MAE), and Root Mean Squared Error (RMSE). Due to the risk of R² inflation, which can occur when including multiple PGSs and lead to an overestimation of explained variance [43], we report the *R^2^*adj. The R²adj accounts for the number of predictors and sample size, providing a more accurate measure of model performance.

To validate our model results further, logistic regression modeled cognitive impairment probability, with model fit and complexity assessed by Akaike Information Criterion (AIC), where a lower AIC indicates better balance. Sensitivity analyses excluded participants with T2D and repeated main analyses to ensure robustness. We didn’t include T2D as a covariate or interaction term in all models because we have PGS_T2D_ in many analyses and including T2D case-control status with PGS_T2D_ would confound the association.

All models included PGS(s) as predictor(s) and adjusted for sex, age, genotyping batch, and 10 ancestry PCs. Models without PGSs excluded genotyping batch and PCs as covariates. All variables, excluding sex, age, and the PCs, were standardized. Analyses were conducted using R version 4.1.2 [44], and the ‘ggplot2’ package was used to visualize the results.

## Results

### Sample Characteristics

In DMS, genotype data have been collected from 8,366 out of 9,187 participants with phenotypic data. After excluding samples with poor genotype call rates, related individuals, duplicate samples, and ancestry outliers (non-European ancestry), 6,896 genotyped individuals remained. Complete data for all phenotype factors were available for 5,244 (76.04%) out of the 6,896 genotyped participants. A flowchart detailing the study sample selection process is provided in **Supplementary Figure 1**. Compared to the included sample, excluded participants showed no significant differences in age and sex distribution, although exhibited variations in other phenotypic characteristics, including education level, LIBRA score, and cognitive function, as shown in **Supplementary Table 1**.

### Pearson Correlations for PGSs

Correlations among all 17 included PGSs are displayed in **Figure 1** (see **Supplementary Table 4** for precise *r* and *p* values). The most significant correlations were found between PGS_BD_ and PGS_SCZ_ (*r* = 0.348, *p* < .001), and between PGS_IQ_ and PGS_EA_ (*r* = 0.274, *p* < .001). There was a correlation between PGS_ASD_ and PGS_ADHD_ (*r* = 0.241, *p* < .001) as well as between the brain structure PGSs PGS_BV_ and PGS_SA_ (*r* = 0.207, *p* < .001). Correlations among the other PGSs were negligible (*r* < 0.2). The low correlations observed suggest a minimal likelihood of multicollinearity, thereby supporting the feasibility of advancing to a multi-PGS linear regression model in subsequent analyses.

**Figure 1.**
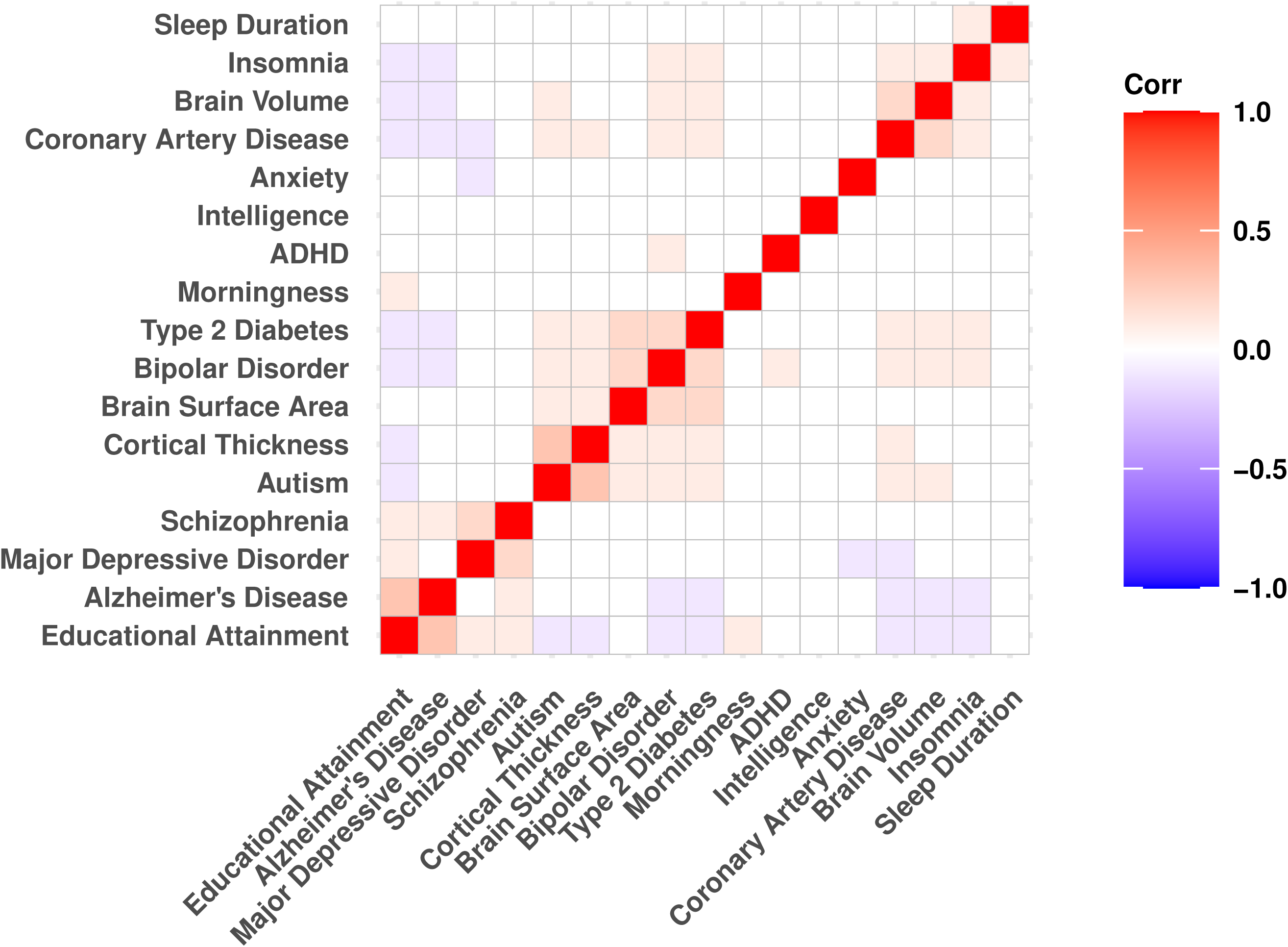
Clustered heat map of Pearson correlations among 17 polygenic scores. Note: ADHD = attention-deficit/hyperactivity disorder. The computed correlation was determined using the Pearson method with listwise deletion and was ordered using the Ward.D2 hierarchical clustering method.

#### Polygenic Score (PGS) Models

##### Preliminary polygenic score analyses

As a validity check, we evaluated the associations of 10 PGSs with available corresponding phenotypes in DMS. All ten demonstrated the expected significant associations with their respective phenotypes (**Supplementary Table 5**).

##### Overall Cognition outcome

Across all single-PGS models, 5 PGSs were identified as significantly associated with overall cognition following FDR correction, detailed in **Table 1**. The PGS_IQ_ showed the strongest association (β, [95% CI], Δ *R*^2^adj) (β = 0.109 [0.094, 0.124], 2.73%). Other PGSs significantly associated with overall cognition included: PGS_EA_ (β = 0.054, [0.039, 0.068], 0.67%), PGS_ADHD_ (β = −0.042, [−0.057, −0.028], 0.40%), PGS_SCZ_ (β = −0.037, [−0.052, −0.022], 0.28%), PGS_CT_ (β = −0.026, [−0.041, −0.011], 0.15%). PGS_BV_, PGS_morningness_, PGS_SA_, PGS_sleepduration_, PGS_insomnia_, PGS_anxiety_, PGS_T2D_ and PGS_CAD_ exhibited nominally significant (uncorrected) associations with overall cognition only. No significant associations with overall cognition were observed for PGS_BD_, PGS_MDD_, PGS_AD_ and PGS_ASD_.

**Table 1.** Associations for 17 polygenic scores (PGSs) with overall cognition in single-PGS and multi-PGS linear regression models.

| PGS<sup>a</sup> | Single PGS Models |  |  |  |  |  | Multi PGS model<sup>b</sup> |  |  |
| --- | --- | --- | --- | --- | --- | --- | --- | --- | --- |
| | p-threshold (best) | $\beta$ | 95% CI | p-value | FDR p-value | Adjusted $\Delta R^{2c}$ | $\beta$ | 95% CI | p-value |
| **Cognition** |  |  |  |  |  |  |  |  |  |
| Intelligence | 0.036 | 0.109 | 0.094; 0.124 | <.001 | <.001 | 2.73% | 0.097 | 0.082; 0.112 | <.001 |
| Educational Attainment | 0.019 | 0.054 | 0.039; 0.068 | <.001 | <.001 | 0.67% | 0.024 | 0.009; 0.039 | 0.002 |
| Alzheimer's Disease | <.001 | -0.011 | -0.025; 0.004 | 0.158 | 1.000 | 0.01% | NA | NA | NA |
| **Mental Health Vulnerability** |  |  |  |  |  |  |  |  |  |
| Major Depressive Disorder | 1.000 | -0.012 | -0.027; 0.003 | 0.114 | 1.000 | 0.02% | NA | NA | NA |
| Anxiety Disorders | 0.010 | -0.018 | -0.033; -0.003 | 0.018 | 0.301 | 0.06% | NA | NA | NA |
| Schizophrenia | 0.042 | -0.037 | -0.052; -0.022 | <.001 | <.001 | 0.28% | -0.026 | -0.041; -0.011 | 0.001 |
| Bipolar Disorder | 0.004 | -0.015 | -0.030; 0 | 0.054 | 0.912 | 0.04% | NA | NA | NA |
| **Neurodevelopmental Vulnerability** |  |  |  |  |  |  |  |  |  |
| ADHD | 0.043 | -0.042 | -0.057; -0.028 | <.001 | <.001 | 0.40% | -0.025 | -0.040; -0.010 | 0.001 |
| Autism Spectrum Disorder | 0.500 | -0.009 | -0.024; 0.006 | 0.237 | 1.000 | 0.01% | NA | NA | NA |
| **Cardiometabolic Vulnerability** |  |  |  |  |  |  |  |  |  |
| Type 2 Diabetes | 0.107 | -0.022 | -0.040; -0.004 | 0.018 | 0.306 | 0.06% | NA | NA | NA |
| Coronary Artery Disease | 0.061 | -0.016 | -0.031; -0.001 | 0.033 | 0.561 | 0.05% | NA | NA | NA |
| **Sleep** |  |  |  |  |  |  |  |  |  |
| Insomnia | 0.053 | -0.018 | -0.033; -0.003 | 0.016 | 0.266 | 0.06% | NA | NA | NA |
| Sleep Duration | 0.002 | 0.018 | 0.004; 0.033 | 0.015 | 0.260 | 0.07% | NA | NA | NA |
| Morningness | 0.005 | 0.020 | 0.005; 0.035 | 0.008 | 0.139 | 0.08% | NA | NA | NA |
| **Brain Structure** |  |  |  |  |  |  |  |  |  |
| Brain Surface Area | 0.009 | 0.019 | 0.005; 0.034 | 0.011 | 0.181 | 0.07% | NA | NA | NA |
| Brain Cortical Thickness | 0.001 | -0.026 | -0.041; -0.011 | 0.001 | 0.009 | 0.15% | -0.023 | -0.037; -0.008 | 0.002 |
| Brain Volume | 0.001 | 0.021 | 0.006; 0.035 | 0.006 | 0.105 | 0.09% | NA | NA | NA |

Note: Abbreviations: $\beta$ , standardized regression coefficient; ADHD, attention-deficit/hyperactivity disorder; FDR, false discovery rate; NA, not applicable; PGS, polygenic score;
<sup>a</sup> All models adjusted for sex, age, genotyping batch and 10 ancestry principal components.
<sup>b</sup> Linear regression model, including all PGSs with FDR $p < .05$ estimated in single-PGS regression models.
<sup>c</sup> Incremental Adjusted $R^2$ of PGS, calculated by Adjusted $R^2$ of full model minus Adjusted $R^2$ of covariate model.

When 5 significant PGSs were simultaneously incorporated into the multi-PGS regression model, its explanatory power improved slightly over that of the most powerful single-PGS model, the PGS_IQ_ (*R*^2^adj: 33.0% for PGS_IQ_ vs. 33.6% for multi-PGS; see **Table 2** and **Supplementary Table 6**). Additionally, no multicollinearity was detected, as evidenced by a VIF < 5 (see **Supplementary Table 7**).

**Table 2.**
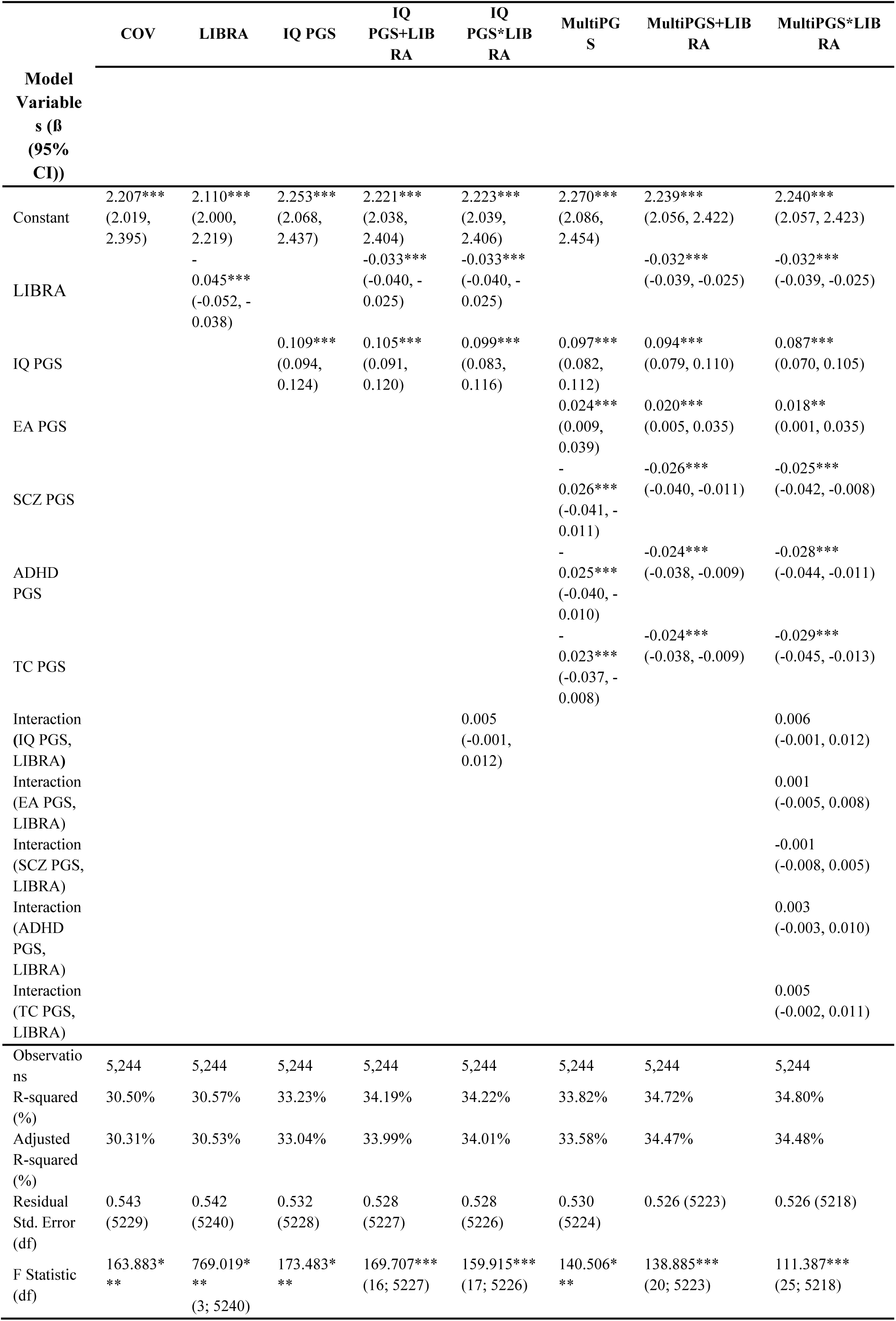
Comparison among the regression models for overall cognition.

##### Specific cognitive domains

Figure 2 illustrates the associations of PGSs with all specific cognitive domains. The majority of the PGSs demonstrate similar association patterns across the three cognitive domains and overall cognition. Notably, the PGS_morningness_ was associated exclusively with memory and not with other cognitive domains, the PGS_BV_ was solely associated with executive function. Following the approach used for overall cognition, we integrated significant FDR-corrected PGSs with specific cognitive domains into the corresponding multi-PGS model, resulting in considerable improvements in *R^2^*adj values across all specific domains. Detailed results for all single and multi-PGS associations across all cognitive domains are presented in **Supplementary Table 6**.

**Figure 2.**
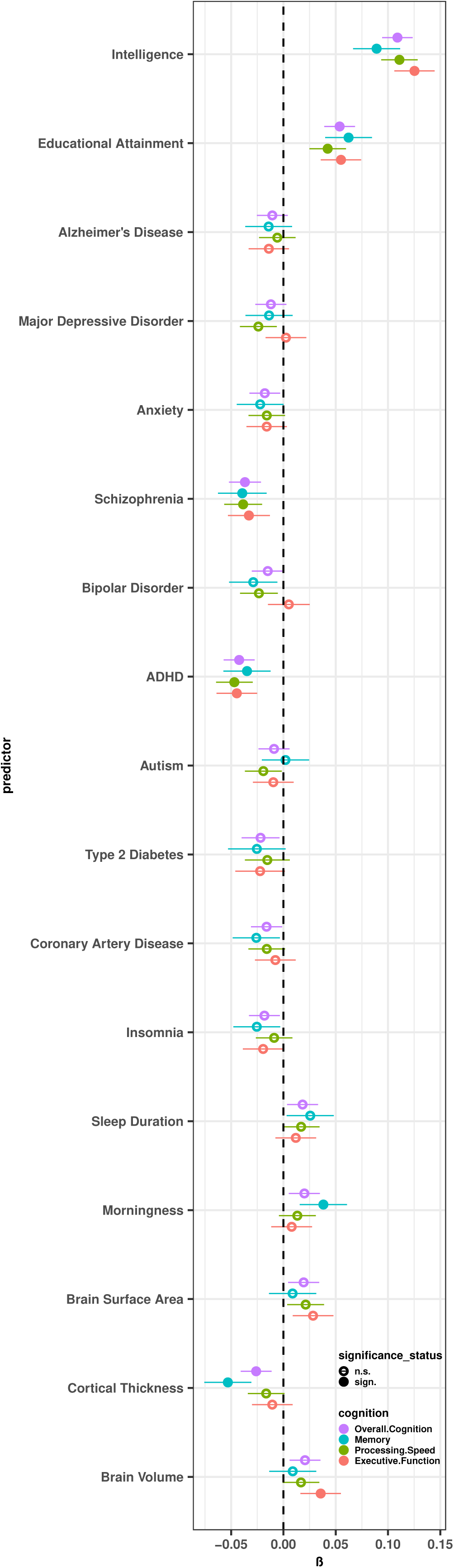
Associations of single polygenic scores (PGSs) with cognition phenotypes. Note: A solid circle (●) denotes a predictor that is significantly (sign.) associated (FDR-adjusted) with this phenotype; an empty circle (○) indicates no significant association (n.s.).

#### The LIBRA Model

The LIBRA score demonstrated a significant association with all cognitive phenotypes (p < 0.001; **Supplementary Table 8**). In terms of overall cognition, the LIBRA score showed significant associative strength (β = −0.045, [−0.052, −0.038], Δ *R*²adj = 2.17%, *R²*adj = 30.53%). In terms of specific domains, LIBRA’s most significant impact was observed in the processing speed domain (β = −0.045, [−0.053, −0.037], Δ *R²*adj = 1.64%, *R²*adj = 25.38%).

#### Comparing the Performance of LIBRA, PGS and Integrated Models

We evaluated LIBRA and genetic models to understand their explanatory performance on all cognition phenotypes. This included assessing the LIBRA model, single PGS models, multi-PGS models, the LIBRA + Single PGS integrated models, and LIBRA + Multi-PGS integrated models. We initially included interaction terms between LIBRA and PGSs in our analysis but found almost no significant interaction effects (see **Table 2** for overall cognition**; Supplementary Table 9** for specific domains). Therefore, our subsequent analysis centered on models excluding these interaction terms.

As presented in **Table 2**, for overall cognition, the *R²*adj values for these models show a progressive increase in explanatory power. The sequence starts from the LIBRA model (*R²*adj = 30.53%), then moves to the most effective single PGS model (IQ; *R²*adj = 33.04%), followed by the multi-PGS models (*R²*adj = 33.58%), and finally, to the combined models of LIBRA + IQ PGS (*R²*adj = 33.99%) and LIBRA + Multi-PGS (*R²*adj = 34.47%). The incremental *R²*adj values, which represent the improvements over the baseline model that includes only covariates, are sequentially 2.17%, 2.73%, 3.27%, 3.68%, and 4.16%, respectively. The same increasing pattern in explanatory power observed for overall cognition is also evident in the three specific cognitive domains (Figure 3).

**Figure 3.**
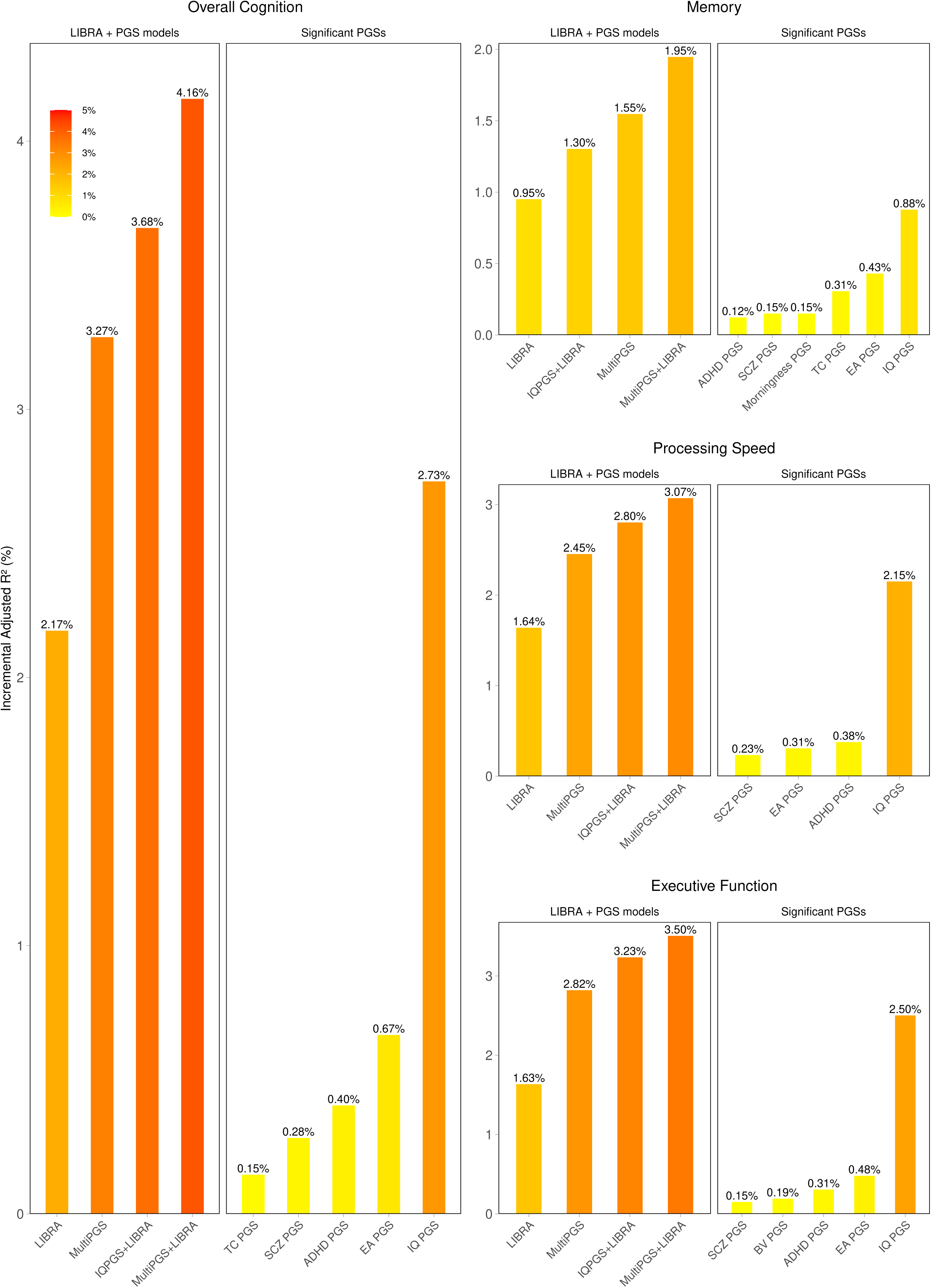
Incremental adjusted R^2^ by PGSs and LIBRA for all cognitive domains. Note: The left panel displays LIBRA and integrated models, the right panel displays all PGS models that are significantly associated with specific domains. The models are arranged from left to right in ascending order of their explanatory power within panels.

A repeated 10-fold cross-validation analysis was conducted to compare the performances of five models across all cognitive phenotypes. This comprehensive analysis consistently highlighted the same patterns across all phenotypes (Figure 4**; Supplementary Table 10**). For overall cognition, the integrated LIBRA + Multi-PGS model surpassed its counterparts by securing the highest mean *R^2^* value, alongside recording the lowest mean MAE and mean RMSE scores. Conversely, the LIBRA model demonstrated the least favorable performance among the evaluated models, registering the lowest mean *R^2^* and the highest mean MAE and mean RMSE scores.

**Figure 4.**
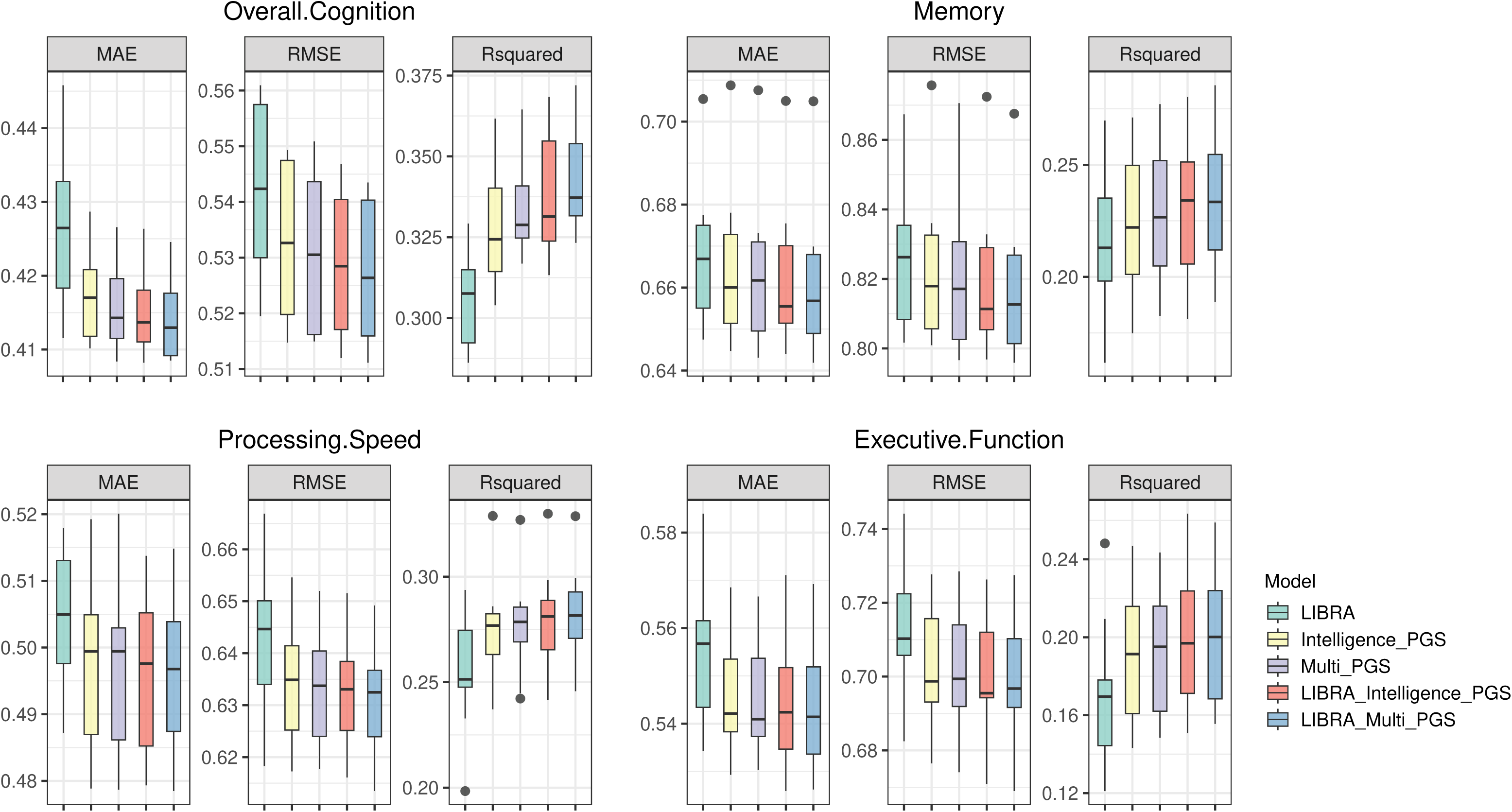
Comparison of model performance based on 10-fold cross-validation. Note: This figure presents the results of 10-fold cross-validation for different models: LIBRA model, Intelligence single-PGS model, Multi-PGS model, a combined LIBRA + Intelligence PGS model and a combined LIBRA + Multi-PGS model. Each model was independently trained and tested 10 times on the same dataset to ensure the reliability of the results. The metrics displayed include Mean Absolute Error (MAE), Root Mean Square Error (RMSE), and R-squared values.

#### Cognitive Impairment

To enhance and substantiate our study results, we used the binary variable ‘cognitive impairment’ as the outcome, defined as significant impairment in any cognitive domain. Utilizing logistic regression analysis, followed by AIC assessment, with the same predictors as those in our linear regression analysis for overall cognition, we aimed to validate the effectiveness of the five models (see **Supplementary Table 11)**. The AIC values indicated the LIBRA + Multi-PGS model, with the lowest AIC at 5448.580, surpassed other models in predicting cognitive impairment (LIBRA: AIC=5507.834; PGS_IQ_: AIC=5474.841; Multi-PGS: AIC=5453.293; LIBRA + PGS_IQ_: AIC=5470.412), aligning closely with our linear regression analysis findings. In the logistic regression, the Multi-PGS model exhibited a marginally better fit compared to the LIBRA + PGS_IQ_ model, marking a slight deviation.

#### Sensitivity Analyses

To confirm the robustness of our findings, we conducted sensitivity analyses by excluding T2D patients and reassessing all associations. The overall association patterns remained consistent, with the same PGSs showing similar associations with overall cognition in the single PGS analysis, with the exception of one additional PGS reaching significance in the non-T2D subset (i.e., morningness) (**Supplementary Table 12**). This indicates these associations are not driven by T2D.

All models retained their significance (*p* < 0.001) at the same level as in the full dataset, both for overall cognition and for all specific domains. The variance explained by the models in the non-T2D subset was similar to that in the full dataset, displaying consistent patterns across models (**Supplementary Table 13**). For the best-fitting model (i.e., multi-PGS + LIBRA), the *R^2^*adj was 0.330 in the non-T2D subset versus 0.345 in the full dataset; this model also demonstrated the smallest AIC value in the logistic regression analysis of cognitive impairment (AIC=4305.495), indicating its optimal fit to the data (**Supplementary Table 14**).

## Discussion

Our primary aim was to integrate single PGS/ multiple PGSs into the established LIBRA score to determine if this combination could enhance the explanation of cognitive function and impairment in older individuals. Across the entire DMS cohort and no-T2D sub-cohort with either continuous traits or binary traits for cognition, our linear and logistic regressions coupled with 10-fold cross-validation analysis revealed a significant enhancement in *R^2^* from integrating PGS into the LIBRA model, whether using the highest-performing single PGS or multiple PGSs. Notably, the addition of multiple PGSs to the LIBRA model yielded the best overall performance. To our knowledge, although many studies have investigated the integration of PGS with phenotypic assessment tools, this paper represents the first to merge single PGS/multiple PGSs with the LIBRA model for cognitive function assessment. Through a thorough evaluation of these score combinations, our study found that while the multi-PGS method is significantly more potent than the single PGS, the most substantial enhancement in predicting cognitive functioning was achieved by incorporating single or multiple PGSs with modifiable lifestyle factors into regression models. This study represents a pioneering effort in combining single/multiple PGSs with the LIBRA model, demonstrating the potential for improved explanatory power and predictive performance in assessing cognitive function and impairment in older individuals.

We found some modest correlations between the 17 PGSs (most correlation coefficients < 0.2). The magnitudes of these correlations are consistent with those reported in a previous study on Pearson correlation estimates between PGSs [25].

In the Single PGS models, five PGSs (IQ, EA, SCZ, ADHD and CT) were significantly associated with overall cognition in the full DMS cohort, aligning with a body of research that focused on association of PGSs with cognitive function [24,45,46,47]. The PGS_IQ_ exhibited the highest explanatory power for cognitive outcomes, significantly surpassing the other PGSs, which is consistent with previous research indicating that PGS_IQ_ provides superior interpretability for cognitive tests compared to PGS_EA_ [48]. Notably, PGS_IQ_ also has a slightly stronger explanatory power compared to LIBRA model in terms of variance. In the non-T2D sub-cohort, the same associations persisted, with the addition of one more significant PGS (i.e., morningness), pointing out that the associations between most PGS and cognition are not driven by T2D. The genetic predisposition to morningness showed a more pronounced cognitive benefit in the absence of T2D, likely due to the diminished impact of metabolic disturbances associated with T2D on brain health. Studies have indicated that metabolic health significantly affects cognitive function. For instance, poor metabolic health, often associated with T2D, has been linked to adverse cognitive outcomes, suggesting that the absence of such disturbances can preserve or enhance cognitive benefits [49].

In three specific cognitive domains, a similar trend was observed. Specifically, PGS_morningness_ and PGS_CT_ were exclusively associated with the memory domain, PGS_BV_ was only related to executive function. Previous GWAS findings for general cognitive ability have identified novel loci associated with brain structure [16], but the associations between structure-related PGS and cognition have rarely been reported. We observed a significant negative relationship between PGS_CT_ and memory, and a positive relationship between PGS_BV_ and executive function, indicating that different aspects of brain anatomy may be subject to unique genetic influences, aligning with prior research [50,51,52,53]. However, the connections between structure-related PGS and cognition extend beyond simple correlations. On the one hand, genetic predispositions determining brain volumes or cortical thicknesses might lead to anatomical changes that impact cognitive functions [54]. On the other hand, it is possible that certain genetic variants simultaneously impact brain structure and specific cognitive functions [55]. Moreover, environmental factors also play a crucial role in shaping both brain anatomy and cognitive abilities, adding another layer of complexity to their relationship. Although we have uncovered some relationships, further detailed studies are needed to understand how these genetic influences shape cognitive trajectories throughout the lifespan. For example, adopting longitudinal designs, integrating multi-omics data, and investigating the interplay between genetic predispositions and environmental factors will provide a comprehensive understanding of these relationships.

For the multi-PGS approach, we combined the significant PGSs after FDR correction into a multi-PGS model [25] and found enhanced explanatory power and prediction ability of multi-PGSs on cognitive function. Our results are in line with a previous study [24], which found that incorporating multiple PGSs improved the explanatory of cognitive ability compared to using a single PGS, suggesting that considering the genetic predisposition of related phenotypes can be an efficient way to increase the model’s performance. This is likely because multiple genetic proxies may account for the interrelated etiology of a phenotype, as opposed to relying on a single genetic proxy. Our study reaffirms that integrating multiple PGSs, including those related to cognitive and non-cognitive traits, can significantly enhance the model’s capacity for explained variance and predictive performance. Overall, the multi-PGS model is a promising method for increasing the ability to explain and predict cognitive dysfunction without requiring individual-level data for the correlated phenotypes.

We integrated PGSs with LIBRA to investigate the performance of a combined model formed by both lifestyle and genotype factors in predicting cognitive function. This approach has been similarly employed in previous research on T2D and CAD [56,57]. Here, we observed that the inclusion of PGSs in LIBRA models significantly improved the models’ predictive power, as indicated by the notable increase in *R²* values from not only the OLS regression but also the average cross-validated performance. These findings imply that the genetic factors captured residual risk (Δ R²adj from 3.46% to 3.94%) that was not quantified by the established lifestyle risk factors. Consequently, the combined model leveraging both genetic and lifestyle factors provides a more robust and accurate prediction of cognitive function in older individuals. Overall, our results showed that the top PGS model and multiple PGS models outperformed the LIBRA-only model and integrating PGSs with the LIBRA score may further optimize prediction accuracy for cognitive abilities.

When examining the relationship between PGSIQ and LIBRA (PGSIQ * LIBRA), we discovered that the R²adj is 34.01%. Interestingly, this value is not significantly different from the combined model (PGSIQ + LIBRA). This trend is also evident in the multi-PGS models, where the R²adj for the multiPGS * LIBRA model closely mirrors that of the multiPGS + LIBRA model. These findings strongly suggest that the interaction effect between PGSIQ and LIBRA does not notably contribute to explaining the variance.

It is also worth noting that the R²adj of 30.31% in the covariate (COV) model demonstrates the significant explanatory power of covariates in our analysis. Among these covariates, age stands out as the most substantial contributor. Age is a pivotal factor affecting cognitive abilities, and controlling for these variables is necessary to ensure the validity and stability of our results. Despite the strong explanatory power of covariates, the contributions of LIBRA and PGS are also considerable and effectively enhance the explanatory power of the model.

Our study has significant strengths. Firstly, our comparison of the predictive power of PGSs with LIBRA scores underscores the clinical value of integrating the LIBRA and multi-PGS model for predicting cognitive outcomes. This comparison is crucial in illustrating the utility of our findings in real-world clinical settings. Secondly, our population-based cohort design enhances the realism and generalizability of our findings, ensuring a more representative sample. Thirdly, by using multiple PGSs to examine their associations with diverse cognitive domains, we provide improved understanding of the role of genetics in cognitive functions. Finally, including binary impaired/unimpaired outcomes in our analysis offers additional insights into the implications of PGSs in cognitive health, which is valuable for future research and potential clinical applications.

There are several limitations worth mentioning. Firstly, our exclusive reliance on cross-sectional data precluded the ability to measure cognitive decline over time. Moreover, the LIBRA score used in the DMS datasets was incomplete, as it did not include cognitive activity, a critical factor for LIBRA calculation, this omission could have led to an underestimation of lifestyle risk. Furthermore, we did not include additional environmental and socioeconomic variables in our models to prevent multicollinearity issues and potential increases in model instability. However, incorporating more environmental and socioeconomic variables could offer a more thorough insight into the factors influencing cognitive risk. Lastly, we use the best-fit P-value threshold in PRSice2 to generate PGSs, which is defined as the threshold at which the PGS is associated with the phenotype (in our study, cognition) and achieves the highest R² in linear regression analysis. However, a potential bias arises because P-values can be influenced by the specific dataset used, which may lead to overfitting. This makes the P-values may be dependent on the particular dataset. Despite conducting internal validation within the database, including repeated 10-fold cross-validation analysis using ‘cognitive impairment’ as a binary outcome variable and performing stratified analysis based on T2D status within the same sample, the lack of external validation with an independent dataset restricts the broader generalizability of our results.

Future research directions may include exploring the relationship between PGSs and the rate of cognitive decline using a longitudinal design, which would provide a better understanding of cognitive aging. Furthermore, considering a broader set of potential confounders, such as disentangling APOE status, is crucial as they may jointly influence both PGS and cognitive function [58]. However, this was beyond the scope of our current study.

In conclusion, our study developed and evaluated a novel approach to better predict cognitive functioning and identify risk factors early on by combining genetic markers (PGS) with risk assessment tools (LIBRA score). This approach provides added value to established clinical risk factors and has potential clinical utility for personalized prevention strategies.

However, it is imperative to acknowledge that, given that LIBRA health and lifestyle factors can be relatively easily obtained from health records, the added benefit and cost-effectiveness of incorporating PGS should be carefully evaluated in real-life screening settings before considering implementation in the general population. Future studies should also validate this approach in other populations and evaluate its clinical utility. Our approach, which combines genomic and phenotypic information, represents a significant step forward in predicting cognitive function, overcoming the limitations of using single PGSs or a single LIBRA score alone.

## Supporting information

Suppemental documents

## Acknowledgments

The Maastricht Study was supported by the European Regional Development Fund via OP-Zuid, the Province of Limburg, the Dutch Ministry of Economic Affairs (grant 31O.041), Stichting De Weijerhorst (Maastricht, the Netherlands), the Pearl String Initiative Diabetes (Amsterdam, the Netherlands), the Cardiovascular Center (CVC, Maastricht, the Netherlands), Mental Health and Neuroscience Research Institute (MHeNS, Maastricht, The Netherlands), Cardiovascular Research Institute Maastricht (CARIM, Maastricht, the Netherlands), Care and Public Health Research Institute (CAPHRI, Maastricht, the Netherlands), Nutrition, Toxicology and Metabolism Research Institute (NUTRIM, Maastricht, the Netherlands), Stichting Annadal (Maastricht, the Netherlands), Health Foundation Limburg (Maastricht, the Netherlands) and by unrestricted grants from Janssen-Cilag B.V. (Tilburg, the Netherlands), Novo Nordisk Farma B.V. (Alphen aan den Rijn, the Netherlands) and Sanofi-Aventis Netherlands B.V. (Gouda, the Netherlands). Author Y. Zhang was supported by the China Scholarship Council (CSC) from the Ministry of Education of P.R. China. Authors S. Guloksuz and B.P.F. Rutten received support from the YOUTH-GEMs project, funded by the European Union’s Horizon Europe program under the grant agreement number: 101057182. S. Guloksuz was supported by the Ophelia research project, ZonMw grant number: 636340001. We declare that there are no conflicts of interest among the authors.

## Data Availability

Participant data are not available for public deposition due to ethical restrictions and privacy regulations. Data from The Maastricht Study can be made available to any researcher who meets the criteria for access to confidential data. Data requests may be submitted to The Maastricht Study Management Team.

## Notes

### Competing Interest Statement

The authors have declared no competing interest.

### Author Declarations

The Medical Ethics Committee of the Maastricht University Medical Center+ (Maastricht UMC+) gave ethical approval for this work, which used data from The Maastricht Study, a population-based cohort study.

## References

1. Frischkorn GT, Schubert AL, Hagemann D. Processing speed, working memory, and executive functions: Independent or inter-related predictors of general intelligence. Intelligence. 2019;75:95–110.

2. Murman DL. The impact of age on cognition. Semin Hear. 2015;36(3):111–121.

3. Johnson W, Deary IJ, McGue M, Christensen K. Genetic and environmental links between cognitive and physical functions in old age. J Gerontol B Psychol Sci Soc Sci. 2009;64(1):65–72.

4. Blokland GAM, Mesholam-Gately RI, Toulopoulou T, Del Re EC, Lam M, DeLisi LE et al. Heritability of neuropsychological measures in schizophrenia and nonpsychiatric populations: A systematic review and meta-analysis. Schizophr Bull. 2017;43(4):788–800.

5. Deary IJ, Spinath FM, Bates TC. Genetics of intelligence. Eur J Hum Genet. 2006;14(6):690–700.

6. Plomin R, Spinath FM. Intelligence: Genetics, genes, and genomics. J Pers Soc Psychol. 2004;86(1):112–129.

7. Bouchard TJ, Jr., McGue M. Familial studies of intelligence: A review. Science. 1981;212(4498):1055–1059.

8. Deckers K, Köhler S, Ngandu T, Antikainen R, Laatikainen T, Soininen H et al. Quantifying dementia prevention potential in the finger randomized controlled trial using the libra prevention index. Alzheimers Dement. 2021;17(7):1205–1212.

9. Rohr S, Pabst A, Baber R, Engel C, Glaesmer H, Hinz A et al. Social determinants and lifestyle factors for brain health: Implications for risk reduction of cognitive decline and dementia. Sci Rep. 2022;12(1):12965.

10. Deckers K, Cadar D, van Boxtel MPJ, Verhey FRJ, Steptoe A, Kohler S. Modifiable risk factors explain socioeconomic inequalities in dementia risk: Evidence from a population-based prospective cohort study. J Alzheimers Dis. 2019;71(2):549–557.

11. Röhr S, Pabst A, Baber R, Engel C, Glaesmer H, Hinz A et al. Social determinants and lifestyle factors for brain health: Implications for risk reduction of cognitive decline and dementia. Sci Rep-Uk. 2022;12(1).

12. Heger IS, Deckers K, Schram MT, Stehouwer CDA, Dagnelie PC, van der Kallen CJH et al. Associations of the lifestyle for brain health index with structural brain changes and cognition: Results from the maastricht study. Neurology. 2021;97(13):e1300–e1312.

13. Anstey KJ, Ee N, Eramudugolla R, Jagger C, Peters R. A systematic review of meta-analyses that evaluate risk factors for dementia to evaluate the quantity, quality, and global representativeness of evidence. J Alzheimers Dis. 2019;70(s1):S165–S186.

14. Schiepers OJG, Kohler S, Deckers K, Irving K, O’Donnell CA, van den Akker M, et al. Lifestyle for brain health (libra): A new model for dementia prevention. Int J Geriatr Psychiatry. 2018;33(1):167–175.

15. Kamboh MI, Fan KH, Yan Q, Beer JC, Snitz BE, Wang X et al. Population-based genome-wide association study of cognitive decline in older adults free of dementia: Identification of a novel locus for the attention domain. Neurobiol Aging. 2019;84:239 e215–239 e224.

16. Davies G, Lam M, Harris SE, Trampush JW, Luciano M, Hill WD et al. Study of 300,486 individuals identifies 148 independent genetic loci influencing general cognitive function. Nat Commun. 2018;9(1):2098.

17. Trampush JW, Yang ML, Yu J, Knowles E, Davies G, Liewald DC et al. Gwas meta-analysis reveals novel loci and genetic correlates for general cognitive function: A report from the cogent consortium. Mol Psychiatry. 2017;22(3):336–345.

18. Davies G, Marioni RE, Liewald DC, Hill WD, Hagenaars SP, Harris SE et al. Genome-wide association study of cognitive functions and educational attainment in uk biobank (n=112 151). Mol Psychiatry. 2016;21(6):758–767.

19. Hagenaars SP, Harris SE, Davies G, Hill WD, Liewald DC, Ritchie SJ et al. Shared genetic aetiology between cognitive functions and physical and mental health in uk biobank (n=112 151) and 24 gwas consortia. Mol Psychiatry. 2016;21(11):1624–1632.

20. Alhola P, Polo-Kantola P. Sleep deprivation: Impact on cognitive performance. Neuropsychiatr Dis Treat. 2007;3(5):553–567.

21. Blackwell T, Yaffe K, Ancoli-Israel S, Redline S, Ensrud KE, Stefanick ML et al. Association of sleep characteristics and cognition in older community-dwelling men: The mros sleep study. Sleep. 2011;34(10):1347–1356.

22. Shi L, Chen SJ, Ma MY, Bao YP, Han Y, Wang YM et al. Sleep disturbances increase the risk of dementia: A systematic review and meta-analysis. Sleep Med Rev. 2018;40:4–16.

23. Ma Y, Liang L, Zheng F, Shi L, Zhong B, Xie W. Association between sleep duration and cognitive decline. JAMA Netw Open. 2020;3(9):e2013573.

24. Krapohl E, Patel H, Newhouse S, Curtis CJ, von Stumm S, Dale PS et al. Multi-polygenic score approach to trait prediction. Mol Psychiatry. 2018;23(5):1368–1374.

25. Schoeler T, Choi SW, Dudbridge F, Baldwin J, Duncan L, Cecil CM et al. Multi-polygenic score approach to identifying individual vulnerabilities associated with the risk of exposure to bullying. JAMA Psychiatry. 2019;76(7):730–738.

26. Schram MT, Sep SJ, van der Kallen CJ, Dagnelie PC, Koster A, Schaper N et al. The maastricht study: An extensive phenotyping study on determinants of type 2 diabetes, its complications and its comorbidities. Eur J Epidemiol. 2014;29(6):439–451.

27. van der Elst W, van Boxtel MP, van Breukelen GJ, Jolles J. The letter digit substitution test: Normative data for 1,858 healthy participants aged 24-81 from the maastricht aging study (maas): Influence of age, education, and sex. J Clin Exp Neuropsychol. 2006;28(6):998–1009.

28. Van der Elst W, Van Boxtel MP, Van Breukelen GJ, Jolles J. The concept shifting test: Adult normative data. Psychol Assess. 2006;18(4):424–432.

29. Van der Elst W, Van Boxtel MP, Van Breukelen GJ, Jolles J. The stroop color-word test: Influence of age, sex, and education; and normative data for a large sample across the adult age range. Assessment. 2006;13(1):62–79.

30. Van der Elst W, van Boxtel MP, van Breukelen GJ, Jolles J. Rey’s verbal learning test: Normative data for 1855 healthy participants aged 24-81 years and the influence of age, sex, education, and mode of presentation. J Int Neuropsychol Soc. 2005;11(3):290–302.

31. Lam M, Awasthi S, Watson HJ, Goldstein J, Panagiotaropoulou G, Trubetskoy V et al. Ricopili: Rapid imputation for consortias pipeline. Bioinformatics. 2020;36(3):930–933.

32. Chang CC, Chow CC, Tellier LC, Vattikuti S, Purcell SM, Lee JJ. Second-generation plink: Rising to the challenge of larger and richer datasets. Gigascience. 2015;4:7.

33. Loh PR, Danecek P, Palamara PF, Fuchsberger C, Reshef YA, Finucane HK et al. Reference-based phasing using the haplotype reference consortium panel. Nature Genetics. 2016;48(11):1443–1448.

34. Das S, Forer L, Schonherr S, Sidore C, Locke AE, Kwong A et al. Next-generation genotype imputation service and methods. Nat Genet. 2016;48(10):1284–1287.

35. Genomes Project C, Auton A, Brooks LD, Durbin RM, Garrison EP, Kang HM et al. A global reference for human genetic variation. Nature. 2015;526(7571):68–74.

36. Choi SW, O’Reilly PF. Prsice-2: Polygenic risk score software for biobank-scale data. Gigascience. 2019;8(7).

37. Lee JJ, Wedow R, Okbay A, Kong E, Maghzian O, Zacher M et al. Gene discovery and polygenic prediction from a genome-wide association study of educational attainment in 1.1 million individuals. Nat Genet. 2018;50(8):1112–1121.

38. Deckers K, van Boxtel MP, Schiepers OJ, de Vugt M, Munoz Sanchez JL, Anstey KJ et al. Target risk factors for dementia prevention: A systematic review and delphi consensus study on the evidence from observational studies. Int J Geriatr Psychiatry. 2015;30(3):234–246.

39. Deckers K, Kohler S, van Boxtel M, Verhey F, Brayne C, Fleming J. Lack of associations between modifiable risk factors and dementia in the very old: Findings from the cambridge city over-75s cohort study. Aging Ment Health. 2018;22(10):1272–1278.

40. Kroenke K, Spitzer RL, Williams JB. The phq-9: Validity of a brief depression severity measure. J Gen Intern Med. 2001;16(9):606–613.

41. Benjamini Y, Hochberg Y. Controlling the false discovery rate - a practical and powerful approach to multiple testing. J R Stat Soc B. 1995;57(1):289–300.

42. O’Brien RM. A caution regarding rules of thumb for variance inflation factors. Qual Quant. 2007;41(5):673–690.

43. O’brien RM. A caution regarding rules of thumb for variance inflation factors. Quality & Quantity. 2007;41(5):673–690.

44. R Core Team. R: A language and environment for statistical computing.. 2021.

45. Ritchie SJ, Hill WD, Marioni RE, Davies G, Hagenaars SP, Harris SE et al. Polygenic predictors of age-related decline in cognitive ability. Mol Psychiatry. 2020;25(10):2584–2598.

46. He Q, Mam-Lam-Fook CJ, Chaignaud J, Danset-Alexandre C, Iftimovici A, Hauguel JG et al. Influence of polygenic risk scores for schizophrenia and resilience on the cognition of individuals at-risk for psychosis. Transl Psychiat. 2021;11(1).

47. Zhang LS, Hill SK, Guo B, Wu BL, Alliey-Rodriguez N, Eum S et al. Impact of polygenic risk for coronary artery disease and cardiovascular medication burden on cognitive impairment in psychotic disorders. Prog Neuro-Psychoph. 2022;113.

48. Genc E, Schluter C, Fraenz C, Arning L, Metzen D, Nguyen HP et al. Polygenic scores for cognitive abilities and their association with different aspects of general intelligence-a deep phenotyping approach. Mol Neurobiol. 2021;58(8):4145–4156.

49. Yun JS, Jung SH, Shivakumar M, Xiao B, Khera AV, Won HH et al. Polygenic risk for type 2 diabetes, lifestyle, metabolic health, and cardiovascular disease: A prospective uk biobank study. Cardiovasc Diabetol. 2022;21(1).

50. Oschwald J, Guye S, Liem F, Rast P, Willis S, Röcke C et al. Brain structure and cognitive ability in healthy aging: A review on longitudinal correlated change. Rev Neuroscience. 2020;31(1):1–57.

51. Deary IJ, Cox SR, Hill WD. Genetic variation, brain, and intelligence differences. Molecular Psychiatry. 2022;27(1):335–353.

52. Thompson PM, Cannon TD, Narr KL, van Erp T, Poutanen VP, Huttunen M et al. Genetic influences on brain structure. Nat Neurosci. 2001;4(12):1253–1258.

53. Plomin R, Kosslyn SM. Genes, brain and cognition. Nat Neurosci. 2001;4(12):1153–1154.

54. Williams CM, Peyre H, Ramus F. Brain volumes, thicknesses, and surface areas as mediators of genetic factors and childhood adversity on intelligence. Cerebral Cortex. 2023;33(10):5885–5895.

55. Brouwer RM, Klein M, Grasby KL, Schnack HG, Jahanshad N, Teeuw J et al. Genetic variants associated with longitudinal changes in brain structure across the lifespan. Nat Neurosci. 2022;25(4):421–432.

56. He Y, Lakhani CM, Rasooly D, Manrai AK, Tzoulaki I, Patel CJ. Comparisons of polyexposure, polygenic, and clinical risk scores in risk prediction of type 2 diabetes. Diabetes Care. 2021;44(4):935–943.

57. Weijmans M, de Bakker PI, van der Graaf Y, Asselbergs FW, Algra A, Jan de Borst G et al. Incremental value of a genetic risk score for the prediction of new vascular events in patients with clinically manifest vascular disease. Atherosclerosis. 2015;239(2):451–458.

58. Wisdom NM, Callahan JL, Hawkins KA. The effects of apolipoprotein e on non-impaired cognitive functioning: A meta-analysis. Neurobiol Aging. 2011;32(1):63–74.

