## Supplementary material for "Comparison of Single Polygenic, Multiple Polygenic Risk, and Lifestyle for Brain Health Index in Explaining Cognitive Function Among Middle-aged and Older Adults in The Maastricht Study": Suppemental documents

### Supplementary Methods

#### Cognitive performance

The composite memory score was derived from the Verbal Learning Test by assigning weightage to total immediate and delayed recall scores. The processing speed domain was evaluated using a combination of tests including Stroop Colour Word Test Part I and II, Concept Shifting Test Part A and B, and Letter-Digit Substitution Test. Executive function and attention were assessed through Stroop Colour Word Test Part III and Concept Shifting Test Part C. Raw test scores were transformed into z-scores and standardized scores of the Stroop Colour Word Test and Concept Shifting Test were inverted to indicate better cognitive performance. The domain-specific scores were calculated by averaging the z-scores obtained from (sub) tests within that domain (e.g., memory = z-score immediate recall + z-score delayed recall / 2).

In the Verbal Learning Test [1] participants were shown a list of 15 unrelated, monosyllabic words on a computer screen in five consecutive trials. After each trial, they were asked to recall as many words as possible in any order. After 20 minutes, they were asked again to recall the words. The outcomes measured were the total number of correctly recalled words across the five trials (total immediate recall) and the number of correctly recalled words during delayed recall.

The Stroop Colour Word Test [2] comprised three sections. Firstly, participants were asked to verbally read out colour names, such as red, blue, yellow, and green, which were printed in black ink (Part I). Secondly, they were instructed to name solid-coloured patches (Part II). In the final section, participants were required to name the ink colour of colour names that were printed in an incongruent colour (e.g., they were asked to say "red" when the word "yellow" was printed in red) (Part III). The time taken to complete Part III was adjusted based on the average time taken to complete Part I and II.

The Concept Shifting Test [3], which was a variation of the Trailing Making Test, consisted of four subtasks. Each subtask presented participants with 16 small circles arranged along a larger imaginary circle. The circles contained digits, letters, or were empty. The instructions for each subtask were to quickly cross-out the digits in ascending order (Part A), the letters in alphabetic order (Part B), and the letters and digits in alternating order (Part C). Additionally, participants were required to cross-out empty circles in a clockwise direction in two consecutive trials (Part 0) to control for basic motor speed. The time taken to complete subtasks A and B was adjusted to account for the average time taken to complete Part 0, while the time taken to complete Part C was adjusted for the average time taken to complete Parts A and B.

The Letter-Digit Substitution Test [4] involved matching digits to letters based on a given key that paired each number from 1 to 9 with a unique letter. The primary measure of interest was the total number of accurate substitutions made within a time limit of 90 seconds.

#### Genotyping and imputation

Genotyping was conducted using the Illumina Infinium Global Screening Array BeadChip at the Human Genotyping Facility of the Genetic Laboratory of the Department of Internal Medicine, Erasmus University Medical Center, Rotterdam, Netherlands, achieving an initial genotyping success rate of 95%. Quality control(QC) and imputation were executed using the Rapid Imputation for COnsortia PipeLIne (RICOPILI) [5].Preliminary QC included verifying sex discrepancies between self-reported and genotype-based data, identifying related or duplicate samples via identity-by-descent estimation, and excluding strand-ambiguous SNPs and duplicate markers, all performed in Plink 1.9 [6].

Subsequent QC measures focused on ensuring data accuracy and reliability. Samples with over 1% missing data and SNPs with a missing rate above 1% were removed. Additionally, SNPs exceeding 4 Mendelian errors and samples with more than 10,000 Mendelian errors were excluded.

Imputation was carried out using the RICOPILI pipeline, involving prephasing with Eagle v2.3.5 [7] and imputation to the 1000 Genomes Phase 3 reference panel [8] with Minimac3 [9] (increasing the number of available SNP genotypes from genotyped to imputed), where genotype dosage data (i.e., based on imputation probabilities) were converted to best guess genotypes (allele pair A/C/T/G format) at a threshold of $p > 0.8$.

Further QC steps post-imputation included identifying heterozygosity outliers (Fhet threshold 0.2), enforcing Hardy-Weinberg equilibrium (threshold $< 1 \times 10^{-6}$), removing monomorphic SNPs, and ensuring a minimum of 10 chromosome X SNPs for sex determination. Before calculating PGSs, we removed individuals who shared a third-degree relative or closer (relatedness coefficient cut-off: 0.125) as PRSice-2 does not allow for relatedness. For PGS, we selected SNPs with an INFO score above 0.1 and a minor allele frequency (MAF) of at least 0.01, and the ancestry background for European ancestry individuals was determined using the first 10 principal components (PCs) in Plink 1.9. All computational processes were conducted in a Linux environment, Ubuntu 18.04.5 LTS release.

#### LIBRA index

The LIBRA factors were derived from clinical data obtained from physical examination or self-reported questionnaires at the baseline measurement of The Maastricht Study and were then dichotomized (presence or absence of the factor) according to established cut-offs [10], except for the diet factor, we replaced the Mediterranean diet with the Dutch Healthy Eating Index and chose 85 as the cut-off. This arbitrary cutoff point was selected as it reflects adherence of around 30% in the current population, a comparable cutoff point that has been used for overall good adherence to the Mediterranean diet for LIBRA calculation in the Maastricht study[10]. The LIBRA total score was calculated by assigning a weight (positive for the presence of risk factors and negative for the presence of protective factors) to each factor based on the relative risks obtained from published meta-analyses [11,12]. These weights were then standardized and summed to obtain a total score. A higher LIBRA score indicates a higher risk of dementia, with scores ranging from -5.9 to 12.7. All LIBRA factors could be operationalized in The Maastricht Study, with the exception of the LIBRA factor high cognitive activity. Data on engagement in cognitively stimulating activities was not available in the dataset, therefore, this LIBRA factor could not be included in the risk calculation. The protective factors that were operationalized in this study were adherence to a Mediterranean diet and low to moderate alcohol use. The risk factors that were operationalized were physical inactivity, smoking, obesity, depression, T2D, hypertension, hypercholesterolemia, heart disease, and chronic kidney disease. A summary of all individual LIBRA factors assigned weights, and operationalization in this dataset can be found in **Supplementary Table 3**.

### Supplementary Tables

#### Supplementary Table 1. Characteristics of the study sample and comparison of participants that were included and excluded

| Characteristic | Excluded, N = 3,943[1] | Included, N = 5,244[1] | p-value[2] |
|---|---|---|---|
| Age | 59 (9) | 60 (8) | 0.8 |
| Sex | | | 0.005 |
| male | 2,029 (51%) | 2,542 (48%) | |
| female | 1,914 (49%) | 2,702 (52%) | |
| Educational level from cognitive test (3 categories) | | | <0.001 |
| low (no education/ primary education/ lower vocational education) | 700 (19%) | 686 (13%) | |
| middle (intermediate vocational education/ higher secondary education/ higher vocational education | 1,570 (43%) | 2,106 (40%) | |
| high (higher professional education/ university education) | 1,372 (38%) | 2,452 (47%) | |
| LIBRA total score | 1.67 (2.23) | 1.23 (2.24) | <0.001 |
| Overall Cognition | -0.06 (0.70) | 0.06 (0.65) | <0.001 |
| Memory | -0.08 (0.99) | 0.06 (0.93) | <0.001 |
| Processing Speed | -0.09 (0.84) | 0.08 (0.75) | <0.001 |
| Executive Attention | -0.07 (0.83) | 0.06 (0.79) | <0.001 |
| Type 2 Diabetes | 1,009 (26%) | 995 (19%) | <0.001 |
| Significant impairment in any cognitive domain (<-1.5 SD below norm) | 1,050 (30%) | 1,153 (22%) | <0.001 |
| Significant Memory impairment (<-1.5 SD below norm) | 404 (12%) | 422 (8.0%) | <0.001 |
| Significant Processing Speed impairment (<-1.5 SD below norm) | 762 (22%) | 807 (15%) | <0.001 |
| Significant Executive/Attention impairment (<-1.5 SD below norm) | 217 (6.2%) | 175 (3.3%) | <0.001 |

[1]Mean (SD); n (%)

[2]Wilcoxon rank sum test; Pearson's Chi-squared test

#### Supplementary Table 2 Overview of GWAS summary statistics used for 17 PGSs

| **Trait** | **Sample size** | **Reference*** |
|---|---|---|
| Diagnosis of depression | 480,359 | Wray NR, Ripke S, Mattheisen M, et al. Genome-wide association analyses identify 44 risk variants and refine the genetic architecture of major depression. Nat Genet. 2018;50(5):668-681. doi:10.1038/s41588-018-0090-3. |
| Alzheimer | 455,258 | Jansen IE, Savage JE, Watanabe K, et al. Genome-wide meta-analysis identifies new loci and functional pathways influencing Alzheimer's disease risk [published correction appears in Nat Genet. 2020 Mar;52(3):354]. Nat Genet. 2019;51(3):404-413. doi:10.1038/s41588-018-0311-9 |
| Schizophrenia | 320,449 | Trubetskoy V, Pardiñas AF, Qi T, et al. Mapping genomic loci implicates genes and synaptic biology in schizophrenia. Nature. 2022;604(7906):502-508. doi:10.1038/s41586-022-04434-5 |
| Intelligence | 269,867 | Savage JE, Jansen PR, Stringer S, et al. Genome-wide association meta-analysis in 269,867 individuals identifies new genetic and functional links to intelligence. Nat Genet. 2018;50(7):912-919. doi:10.1038/s41588-018-0152-6. |
| Educational attainment | 3,037,499 | Okbay A, Wu Y, Wang N, Jayashankar H, Bennett M, Nehzati SM, Sidorenko J, Kweon H, Goldman G, Gjorgjieva T, Jiang Y. Polygenic prediction of educational attainment within and between families from genome-wide association analyses in 3 million individuals. Nature genetics. 2022 Apr;54(4):437-49. |
| Brain cortical surface area | 51,665 | Grasby KL, Jahanshad N, Painter JN, et al. The genetic architecture of the human cerebral cortex [published correction appears in Science. 2021 Oct 22;374(6566):eabm7211]. Science. 2020;367(6484):eaay6690. doi:10.1126/science.aay6690 |
| Brain cortical thickness | 51,665 | Grasby KL, Jahanshad N, Painter JN, et al. The genetic architecture of the human cerebral cortex [published correction appears in Science. 2021 Oct 22;374(6566):eabm7211]. Science. 2020;367(6484):eaay6690. doi:10.1126/science.aay6690 |
| Brain volume | 47,316 | Jansen, P.R., Nagel, M., Watanabe, K. et al. Genome-wide meta-analysis of brain volume identifies genomic loci and genes shared with intelligence. Nat Commun 11, 5606 (2020). https://doi.org/10.1038/s41467-020-19378-5 |
| Insomnia | 1,331,010 | Jansen PR, Watanabe K, Stringer S, et al. Genome-wide analysis of insomnia in 1,331,010 individuals identifies new risk loci and functional pathways. Nat Genet. 2019;51(3):394-403. doi:10.1038/s41588-018-0333-3 |
| Sleep duration | 384,317 | |
| Morningness | 345,552 | |
| Type 2 diabetes | 251,740 | Mahajan A, Spracklen CN, Zhang W, et al. Multi-ancestry genetic study of type 2 diabetes highlights the power of diverse populations for discovery and translation. Nat Genet. 2022;54(5):560-572. doi:10.1038/s41588-022-01058-3 |
| Attention deficit hyperactivity disorder. | 55,374 | Demontis D, Walters RK, Martin J, et al. Discovery of the first genome-wide significant risk loci for attention deficit/hyperactivity disorder. Nat Genet. 2019;51(1):63-75. doi:10.1038/s41588-018-0269-7 |
| Anxiety | 83,566 | Purves KL, Coleman JRI, Meier SM, et al. A major role for common genetic variation in anxiety disorders. Mol Psychiatry. 2020;25(12):3292-3303. doi:10.1038/s41380-019-0559-1 |
| Coronary artery disease | 250,736 | Van Der Harst, P., & Verweij, N. (2018). Identification of 64 novel genetic loci provides an expanded view on the genetic architecture of coronary artery disease. Circulation research, 122(3), 433-443. |
| Bipolar disorder | 413,466 | Mullins N, Forstner AJ, O'Connell KS, et al. Genome-wide association study of more than 40,000 bipolar disorder cases provides new insights into the underlying biology. Nat Genet. 2021;53(6):817-829. doi:10.1038/s41588-021-00857-4 |
| Autism | 46,350 | Grove J, Ripke S, Als TD, et al. Identification of common genetic risk variants for autism spectrum disorder. Nat Genet. 2019;51(3):431-444. doi:10.1038/s41588-019-0344-8 |

* All GWAS summary statistics used for polygenic scoring are based on European ancestry samples.

**Supplementary Table 3. Operationalization of LIBRA factors in The Maastricht Study**

| LIBRA factor | Weight | Operationalization in The Maastricht Study |
|---|---|---|
| Adherence to a healthy diet | -1.7 | The Dutch Healthy Diet index 2015 minus alcohol component (range 0-130). Scores ≥85 are categorized as adherence to the diet. |
| Physical inactivity | 1.1 | <150 min/wk of (self-reported on CHAMPS questionnaire) moderate to vigorous physical activity in the past 2 week was categorized as physically inactive. |
| Smoking | 1.5 | Self-reported data on smoking cigarettes based on an item of the FFQ. Current smokers were included in the risk score. |
| Low to moderate alcohol intake | -1 | Self-reported alcohol intake based on the FFQ. Low to moderate alcohol use was defined as <70 g/wk. |
| Obesity | 1.6 | BMI ≥30 kg/m2 calculated from physical examination at the research center. |
| Depression | 2.1 | Current major or minor depressive episode based on the MINI or presence of moderate to severe depressive symptoms based on the PHQ9 (range 0–27; cutoff ≥10). |
| Type 2 diabetes | 1.3 | Glucose tolerance status based on fasting glucose (≥7.0), oral glucose tolerance test (≥11.1), or information on current diabetes medications. |
| Hypertension | 1.6 | Average systolic blood pressure ≥140 mm Hg, diastolic blood pressure ≥90, or current antihypertensive medication use. |
| High cholesterol | 1.4 | Serum total cholesterol ≥6.5 mmol/L. |
| Heart disease | 1 | Self-reported history of cardiovascular disease (cerebrovascular accidents excluded). |
| Chronic kidney disease | 1.1 | Levels of serum cystatin C of <60 and/or average albuminuria categories, based on average urinary albumin excretion. Microalbuminuria and macro albuminuria were defined as risk. |
| Cognitive activity | -3.2 | Data not available in dataset. |

#### Supplementary Table 4. Pearson correlations among all 17 included polygenic scores (PGSs)

| | Intelligence | Educational Attainment | Alzheimer's Disease | Major Depressive Disorder | Anxiety | Schizophrenia | Bipolar Disorder | ADHD | Autism | Type 2 Diabetes | Coronary Artery Disease | Insomnia | Sleep Duration | Morningness | Brain Surface Area | Cortical Thickness | Brain Volume |
|---|---|---|---|---|---|---|---|---|---|---|---|---|---|---|---|---|---|
| **Intelligence** | 1 | | | | | | | | | | | | | | | | |
| **Educational Attainment** | 0.004 *(.796)* | 1 | | | | | | | | | | | | | | | |
| **Alzheimer's Disease** | 0.007 *(.599)* | **0.274 (<.001)** | 1 | | | | | | | | | | | | | | |
| **Major Depressive Disorder** | -0.013 *(.346)* | 0.087 *(<.001)* | 0.049 *(<.001)* | 1 | | | | | | | | | | | | | |
| **Anxiety** | -0.015 *(.278)* | -0.029 *(.039)* | 0.011 *(.412)* | -0.064 *(<.001)* | 1 | | | | | | | | | | | | |
| **Schizophrenia** | -0.015 *(.286)* | 0.075 *(<.001)* | 0.051 *(<.001)* | **0.207 *(<.001)*** | -0.010 *(.465)* | 1 | | | | | | | | | | | |
| **Bipolar Disorder** | 0.013 *(.358)* | -0.061 *(<.001)* | -0.073 *(<.001)* | -0.026 *(.063)* | -0.036 *(.010)* | -0.020 *(.151)* | 1 | | | | | | | | | | |
| **ADHD** | 0.012 *(.387)* | -0.016 *(.247)* | -0.016 *(.255)* | 0.000 *(.996)* | -0.029 *(.033)* | -0.007 *(.633)* | 0.059 *(<.001)* | 1 | | | | | | | | | |
| **Autism** | 0.007 *(.635)* | -0.110 *(<.001)* | -0.007 *(.600)* | -0.038 *(.006)* | 0.024 *(.085)* | -0.024 *(.083)* | 0.138 *(<.001)* | 0.045 *(.001)* | 1 | | | | | | | | |
| **Type 2 Diabetes** | 0.039 *(.005)* | -0.125 *(<.001)* | -0.146 *(<.001)* | -0.042 *(.002)* | 0.015 *(.290)* | -0.011 *(.407)* | 0.160 *(<.001)* | 0.012 *(.403)* | 0.081 *(<.001)* | 1 | | | | | | | |
| **Coronary Artery Disease** | -0.013 *(.344)* | -0.131 *(<.001)* | -0.098 *(<.001)* | -0.073 *(<.001)* | 0.013 *(.339)* | -0.028 *(.039)* | 0.113 *(<.001)* | 0.033 *(.016)* | 0.106 *(<.001)* | 0.102 *(<.001)* | 1 | | | | | | |
| **Insomnia** | 0.012 *(.403)* | -0.069 *(<.001)* | -0.084 *(<.001)* | -0.018 *(.203)* | -0.025 *(.068)* | -0.020 *(.151)* | 0.091 *(<.001)* | 0.046 *(.001)* | 0.014 *(.307)* | 0.059 *(<.001)* | 0.059 *(<.001)* | 1 | | | | | |
| **Sleep Duration** | -0.011 *(.438)* | 0.028 *(.041)* | -0.022 *(.104)* | -0.022 *(.112)* | -0.016 *(.243)* | -0.016 *(.237)* | 0.013 *(.333)* | 0.006 *(.645)* | -0.025 *(.071)* | 0.018 *(.198)* | -0.022 *(.113)* | 0.128 *(<.001)* | 1 | | | | |
| **Morningness** | -0.032 *(.022)* | 0.071 *(<.001)* | 0.034 *(.014)* | -0.005 *(.717)* | -0.007 *(.632)* | -0.027 *(.047)* | 0.046 *(.001)* | 0.002 *(.913)* | 0.031 *(.023)* | -0.015 *(.272)* | -0.037 *(.007)* | 0.002 *(.862)* | -0.008 *(.571)* | 1 | | | |
| **Brain Surface Area** | 0.017 *(.217)* | 0.021 *(.122)* | 0.020 *(.147)* | 0.015 *(.273)* | -0.025 *(.066)* | -0.005 *(.730)* | 0.160 *(<.001)* | 0.020 *(.145)* | 0.086 *(<.001)* | **0.241 *(<.001)*** | -0.016 *(.234)* | 0.015 *(.271)* | 0.022 *(.113)* | 0.034 *(.014)* | 1 | | |
| **Cortical Thickness** | -0.014 *(.302)* | -0.052 *(<.001)* | 0.045 *(.001)* | -0.024 *(.085)* | 0.027 *(.053)* | -0.006 *(.669)* | 0.123 *(<.001)* | 0.034 *(.013)* | **0.348 *(<.001)*** | 0.054 *(<.001)* | 0.068 *(<.001)* | 0.023 *(.093)* | -0.012 *(.375)* | -0.001 *(.959)* | 0.051 *(<.001)* | 1 | |
| **Brain Volume** | 0.029 *(.035)* | -0.064 *(<.001)* | -0.096 *(<.001)* | -0.034 *(.013)* | -0.009 *(.512)* | -0.042 *(.003)* | 0.072 *(<.001)* | 0.050 *(<.001)* | 0.052 *(<.001)* | 0.093 *(<.001)* | 0.183 *(<.001)* | 0.063 *(<.001)* | 0.007 *(.591)* | 0.000 *(.989)* | 0.005 *(.739)* | 0.030 *(.029)* | 1 |

Note: Computed correlation used Pearson-method with listwise-deletion. Values of correlation coefficient > 0.2 are highlighted in bold.

**Supplementary Table 5. Variance explained by each polygenic score in corresponding outcome variables.**

| Polygenic score | Outcome variable | Sample Size | β | SE | p-value | % variance explained |
|---|---|---|---|---|---|---|
| Educational Attainment | Education Level† | 5244 | 0.414 | 0.033 | 4.08E-35 | 10.23% |
| Intelligence | Overall Cognition | 5244 | 0.107 | 0.007 | 1.10E-47 | 31.48% |
| Alzheimer's Disease | Cognitive impairment | 5244 | 0.076 | 0.033 | 2.08E-02 | 0.57% |
| MDD | PHQ-9 score | 5191 | 0.183 | 0.044 | 3.78E-05 | 3.86% |
| Anxiety | The use of anxiolytic drugs† | 5243 | 0.198 | 0.092 | 3.16E-02 | 2.92% |
| Coronary Artery Disease | Cardiovascular disease† | 5244 | 0.332 | 0.043 | 1.09E-14 | 5.80% |
| ADHD | Executive attention | 5244 | -0.052 | 0.01 | 6.17E-08 | 16.02% |
| Sleep Duration | Sleep time | 2987 | -5.525 | 1.458 | 1.54E-04 | 1.90% |
| Insomnia | Fall asleep well† | 5191 | 0.143 | 0.037 | 9.61E-05 | 5.50% |
| Type 2 Diabetes | Diabetes status† | 5244 | 0.496 | 0.047 | 6.93E-26 | 10.95% |

Note: MDD=Major Depressive Disorder, ADHD= attention deficit/hyperactivity disorder. Standardized regression coefficients (β), SEs, and p-values are from general linear (continuous outcomes) or generalized linear (categorical outcomes, indicated with the † symbol) regression models adjusting for age at the time of measuring/reporting the outcome variable, sex, genotyping batch, and 10 ancestry principal components. For continuous outcome variables, the % variance explained is derived from the $R^2$ of the whole model. For categorical outcome variables, the % variance explained is derived from Nagelkerke's $R^2$ of the whole model.

#### Supplementary Table 6. Single-PGS and multi-PGS linear regression models for individual cognitive domains

See SupplTable6_single_multiPGS_cognition.xlsx

#### Supplementary Table 7. Variance Inflation Factors (VIFs) from multi-PGS linear regression models for individual cognitive domains

See SupplTable7_vif_results.xlsx

#### Supplementary Table 8. Multiple linear regression analyses of the association between LIBRA score and cognitive phenotypes

| **Cognition** | **β** | **SE** | **95% CI** | ***p*** | **Model $R^2$** | **Model Adj $R^2$** | **LIBRA Partial $R^2$** | **LIBRA Partial Adj $R^2$** |
|---|---|---|---|---|---|---|---|---|
| Overall Cognition | -0.045 | 0.004 | -0.052; -0.038 | <0.001 | 30.57% | 30.53% | 2.19% | 2.17% |
| Memory | -0.043 | 0.005 | -0.054; -0.033 | <0.001 | 21.50% | 21.46% | 0.97% | 0.95% |
| Processing Speed | -0.045 | 0.004 | -0.053; -0.037 | <0.001 | 25.42% | 25.38% | 1.65% | 1.64% |
| Executive Function | -0.047 | 0.005 | -0.056; -0.038 | <0.001 | 16.95% | 16.90% | 1.65% | 1.63% |

Abbreviations: CI = confidence interval; LIBRA = Lifestyle for Brain Health, Adj $R^2$ = Adjusted $R^2$. The models were adjusted for variables including sex, age, genotyping batch, and ancestry principal components. The total number of observations in the analysis is N=5244.

#### Supplementary Table 9. Comparison among the linear regression models for individual cognitive domains.

| | Dependent variable: Memory | | | | | | | |
|---|---|---|---|---|---|---|---|---|
| **Model Variables (ß (95% CI))** | **COV** | **LIBRA** | **IQPGS** | **IQPGS+LIBRA** | **IQPGS*LIBRA** | **MultiPGS** | **MultiPGS+LIBRA** | **MultiPGS*LIBRA** |
| Constant | 1.961***<br>(1.675, 2.246) | 1.709***<br>(1.542, 1.876) | 1.998***<br>(1.714, 2.282) | 1.967***<br>(1.683, 2.251) | 1.966***<br>(1.682, 2.250) | 2.020***<br>(1.736, 2.303) | 1.989***<br>(1.706, 2.273) | 1.987***<br>(1.703, 2.270) |
| LIBRA | | -0.043***<br>(-0.054, -0.033) | | -0.032***<br>(-0.043, -0.020) | -0.032***<br>(-0.043, -0.020) | | -0.031***<br>(-0.042, -0.020) | -0.031***<br>(-0.042, -0.019) |
| IQ PGS | | | 0.089***<br>(0.067, 0.112) | 0.086***<br>(0.063, 0.108) | 0.090***<br>(0.064, 0.115) | 0.070***<br>(0.046, 0.093) | 0.068***<br>(0.044, 0.091) | 0.072***<br>(0.045, 0.099) |
| EA PGS | | | | | | 0.040***<br>(0.016, 0.063) | 0.036***<br>(0.013, 0.060) | 0.033**<br>(0.007, 0.059) |
| SCZ PGS | | | | | | -0.032***<br>(-0.055, -0.009) | -0.031***<br>(-0.054, -0.008) | -0.025*<br>(-0.051, 0.002) |
| ADHD PGS | | | | | | -0.017<br>(-0.040, 0.005) | -0.016<br>(-0.038, 0.007) | -0.019<br>(-0.045, 0.006) |
| Morningness PGS | | | | | | 0.033***<br>(0.011, 0.056) | 0.034***<br>(0.011, 0.056) | 0.043***<br>(0.017, 0.068) |
| TC PGS | | | | | | -0.051***<br>(-0.073, -0.028) | -0.051***<br>(-0.074, -0.029) | -0.055***<br>(-0.080, -0.030) |
| Interaction (IQ PGS, LIBRA) | | | | | -0.003<br>(-0.013, 0.007) | | | -0.003<br>(-0.014, 0.007) |
| Interaction (EA PGS, LIBRA) | | | | | | | | 0.002<br>(-0.008, 0.013) |
| Interaction (SCZ PGS, LIBRA) | | | | | | | | -0.005<br>(-0.015, 0.005) |
| Interaction (ADHD PGS, LIBRA) | | | | | | | | 0.003<br>(-0.007, 0.013) |
| Interaction (Morningness PGS, LIBRA) | | | | | | | | -0.006<br>(-0.016, 0.003) |
| Interaction (TC PGS, LIBRA) | | | | | | | | 0.003<br>(-0.007, 0.013) |
| Observations | 5,244 | 5,244 | 5,244 | 5,244 | 5,244 | 5,244 | 5,244 | 5,244 |
| R-squared (%) | 21.82% | 21.50% | 22.71% | 23.15% | 23.16% | 23.45% | 23.86% | 23.92% |
| Adjusted R-squared (%) | 21.61% | 21.46% | 22.49% | 22.91% | 22.91% | 23.16% | 23.56% | 23.53% |
| Residual Std. Error | 0.825<br>(5229) | 0.826<br>(5240) | 0.821<br>(5228) | 0.818<br>(5227) | 0.818<br>(5226) | 0.817<br>(5223) | 0.815<br>(5222) | 0.815<br>(5216) |
| F Statistic (df) | 104.243***<br>(14; 5229) | 478.448***<br>(3; 5240) | 102.406***<br>(15; 5228) | 98.402***<br>(16; 5227) | 92.630***<br>(17; 5226) | 80.003***<br>(20; 5223) | 77.939***<br>(21; 5222) | 60.738***<br>(27; 5216) |
| | **Dependent variable: Processing Speed** | | | | | | | |
| **Model Variables (ß (95% CI))** | **COV** | **LIBRA** | **IQPGS** | **IQPGS+LIBRA** | **IQPGS*LIBRA** | **MultiPGS** | **MultiPGS+LIBRA** | **MultiPGS*LIBRA** |
| Constant | 2.328***<br>(2.105, 2.551) | 2.418***<br>(2.288, 2.548) | 2.374***<br>(2.154, 2.594) | 2.344***<br>(2.125, 2.563) | 2.346***<br>(2.127, 2.565) | 2.396***<br>(2.177, 2.616) | 2.367***<br>(2.148, 2.586) | 2.368***<br>(2.149, 2.587) |
| LIBRA | | -0.045***<br>(-0.053, -0.037) | | -0.031***<br>(-0.040, -0.023) | -0.031***<br>(-0.040, -0.023) | | -0.031***<br>(-0.039, -0.022) | -0.030***<br>(-0.039, -0.022) |
| IQ PGS | | | 0.111***<br>(0.094, 0.128) | 0.107***<br>(0.090, 0.125) | 0.100***<br>(0.080, 0.119) | 0.102***<br>(0.084, 0.121) | 0.100***<br>(0.082, 0.118) | 0.091***<br>(0.071, 0.112) |
| EA PGS | | | | | | 0.009<br>(-0.009, 0.027) | 0.006<br>(-0.012, 0.024) | 0.003<br>(-0.017, 0.024) |
| SCZ PGS | | | | | | -0.028***<br>(-0.046, -0.010) | -0.027***<br>(-0.045, -0.009) | -0.030***<br>(-0.050, -0.010) |
| ADHD PGS | | | | | | -0.031*** | -0.030*** | -0.034*** |

| | | | | | | | | |
|---|---|---|---|---|---|---|---|---|
| | | | | | | (-0.049, -0.014) | (-0.047, -0.012) | (-0.053, -0.014) |
| Interaction **(**IQ PGS, LIBRA**)** | | | | | 0.006 (-0.001, 0.014) | | | 0.007* (-0.001, 0.015) |
| Interaction (EA PGS, LIBRA) | | | | | | | | 0.002 (-0.006, 0.010) |
| Interaction (SCZ PGS, LIBRA) | | | | | | | | 0.002 (-0.006, 0.010) |
| Interaction (ADHD PGS, LIBRA) | | | | | | | | 0.003 (-0.005, 0.011) |
| Observations | 5,244 | 5,244 | 5,244 | 5,244 | 5,244 | 5,244 | 5,244 | 5,244 |
| R-squared (%) | 25.59% | 25.42% | 27.75% | 28.41% | 28.45% | 28.09% | 28.72% | 28.78% |
| Adjusted R-squared (%) | 25.39% | 25.38% | 27.54% | 28.19% | 28.21% | 27.84% | 28.46% | 28.46% |
| Residual Std. Error | 0.643 (5229) | 0.643 (5240) | 0.634 (5228) | 0.631 (5227) | 0.631 (5226) | 0.633 (5225) | 0.630 (5224) | 0.630 (5220) |
| F Statistic (df) | 128.435*** (14; 5229) | 595.346*** (3; 5240) | 133.843*** (15; 5228) | 129.652*** (16; 5227) | 122.220*** (17; 5226) | 113.393*** (18; 5225) | 110.776*** (19; 5224) | 91.703*** (23; 5220) |
| | **Dependent variable: Executive Function** | | | | | | | |
| **Model Variables (ß (95% CI))** | **COV** | **LIBRA** | **IQPGS** | **IQPGS+LIBRA** | **IQPGS*LIBRA** | **MultiPGS** | **MultiPGS+LIBRA** | **MultiPGS*LIBRA** |
| Constant | 2.292*** (2.045, 2.539) | 2.174*** (2.031, 2.318) | 2.344*** (2.101, 2.587) | 2.311*** (2.069, 2.553) | 2.315*** (2.073, 2.557) | 2.370*** (2.127, 2.613) | 2.337*** (2.095, 2.579) | 2.340*** (2.098, 2.582) |
| LIBRA | | -0.047*** (-0.056, -0.038) | | -0.035*** (-0.045, -0.025) | -0.035*** (-0.045, -0.025) | | -0.034*** (-0.043, -0.024) | -0.034*** (-0.043, -0.024) |
| IQ PGS | | | 0.125*** (0.106, 0.145) | 0.121*** (0.102, 0.141) | 0.106*** (0.084, 0.128) | 0.113*** (0.093, 0.134) | 0.111*** (0.091, 0.131) | 0.094*** (0.071, 0.117) |
| EA PGS | | | | | | 0.019* (-0.001, 0.038) | 0.015 (-0.005, 0.035) | 0.015 (-0.008, 0.037) |
| SCZ PGS | | | | | | -0.021** (-0.041, -0.002) | -0.021** (-0.041, -0.001) | -0.024** (-0.047, -0.002) |
| ADHD PGS | | | | | | -0.026*** (-0.046, -0.007) | -0.025** (-0.044, -0.005) | -0.030*** (-0.052, -0.008) |
| BV PGS | | | | | | 0.026*** (0.007, 0.045) | 0.026*** (0.007, 0.045) | 0.019* (-0.002, 0.041) |
| Interaction **(**IQ PGS, LIBRA**)** | | | | | 0.013*** (0.004, 0.021) | | | 0.013*** (0.004, 0.022) |
| Interaction (EA PGS, LIBRA) | | | | | | | | 0.0003 (-0.008, 0.009) |
| Interaction (SCZ PGS, LIBRA) | | | | | | | | 0.002 (-0.007, 0.010) |
| Interaction (ADHD PGS, LIBRA) | | | | | | | | 0.004 (-0.005, 0.013) |
| Interaction (BV PGS, LIBRA) | | | | | | | | 0.005 (-0.004, 0.013) |
| Observations | 5,244 | 5,244 | 5,244 | 5,244 | 5,244 | 5,244 | 5,244 | 5,244 |
| R-squared (%) | 16.96% | 16.95% | 19.47% | 20.22% | 20.35% | 19.85% | 20.55% | 20.72% |
| Adjusted R-squared (%) | 16.74% | 16.90% | 19.24% | 19.97% | 20.09% | 19.56% | 20.24% | 20.34% |
| Residual Std. Error | 0.712 (5229) | 0.711 (5240) | 0.701 (5228) | 0.698 (5227) | 0.698 (5226) | 0.700 (5224) | 0.697 (5223) | 0.696 (5218) |
| F Statistic (df) | 76.286*** (14; 5229) | 356.444*** (3; 5240) | 84.271*** (15; 5228) | 82.779*** (16; 5227) | 78.538*** (17; 5226) | 68.088*** (19; 5224) | 67.536*** (20; 5223) | 54.557*** (25; 5218) |

Note: COV = The baseline model that only includes covariates. The asterisks represent significance levels: **p<0.05; ***p<0.01. Models adjusted for age, sex, genotyping batch, and ancestry principal components (PC1-PC10); LIBRA model for age and sex only.

**Supplementary Table 10. 10-fold cross validation for all cognitive domains**
See SupplTable10_ crossvalidation_results.xlsx

**Supplementary Table 11. Validation logistic regression for cognitive impairment**
See SupplTable11_CogIMPAIR_Model_Results.xlsx

**Supplementary Table 12. Sensitivity linear regression for single PGSs and multi-PGS models in the non-T2D subset.**
See SupplTable12_single_multi_allcognition_nonT2D.xlsx

**Supplementary Table 13. Sensitivity linear regression models for individual cognitive domains in the non-T2D subset.**
See SupplTable13_comparison_5regression_models_nonT2D.xlsx

**Supplementary Table 14. Sensitivity logistic regression models for cognitive impairment in the non-T2D subset.**
See SupplTable14_comparison_5logistic_models_nonT2D.xlsx

### Supplementary Figures

#### Supplementary Figure 1. The study population selection process

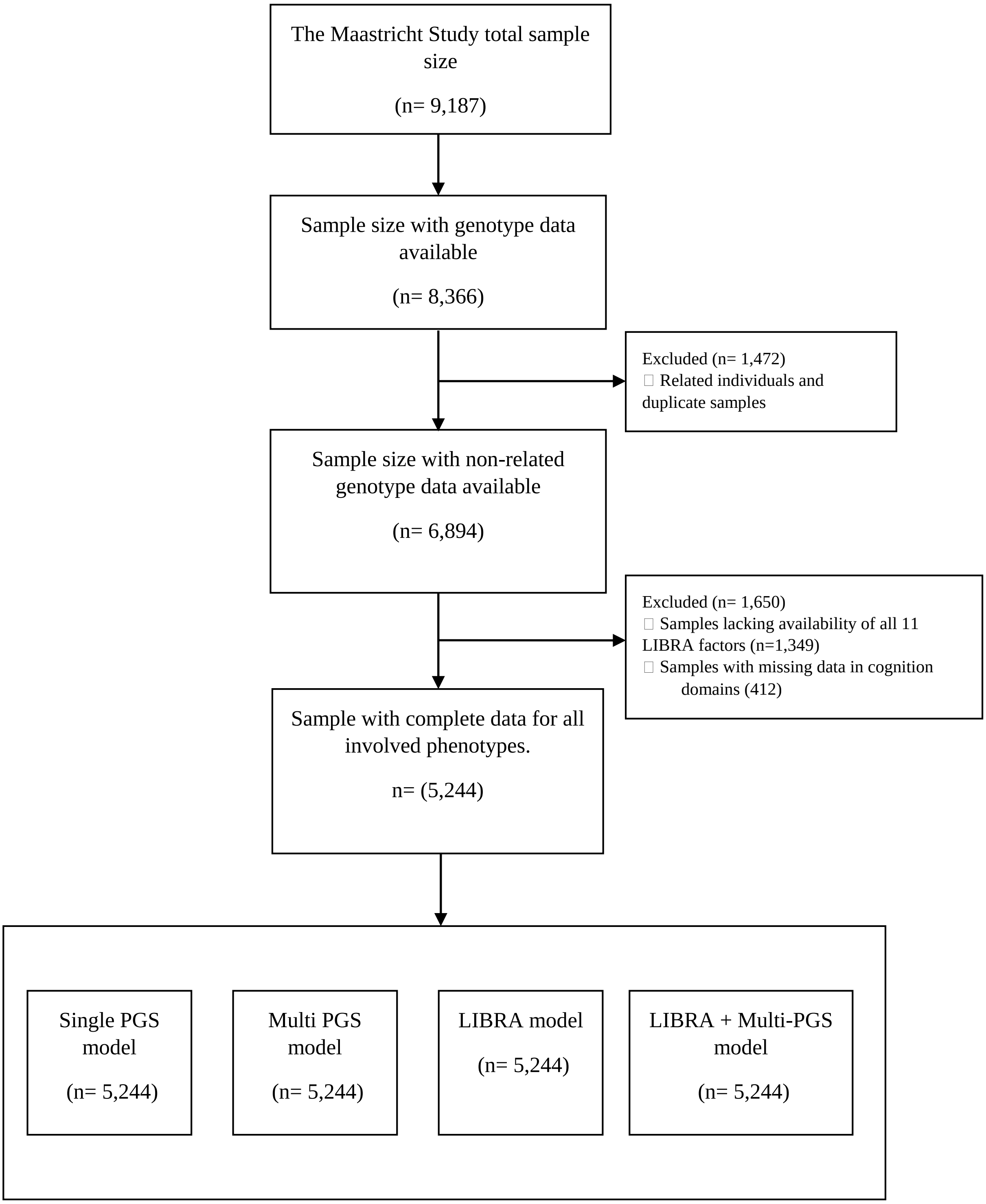

#### References

1. Van der Elst W, van Boxtel MP, van Breukelen GJ, Jolles J. Rey's verbal learning test: Normative data for 1855 healthy participants aged 24-81 years and the influence of age, sex, education, and mode of presentation. J Int Neuropsychol Soc. 2005;11(3):290-302.
2. Van der Elst W, Van Boxtel MP, Van Breukelen GJ, Jolles J. The stroop color-word test: Influence of age, sex, and education; and normative data for a large sample across the adult age range. Assessment. 2006;13(1):62-79.
3. Van der Elst W, Van Boxtel MP, Van Breukelen GJ, Jolles J. The concept shifting test: Adult normative data. Psychol Assess. 2006;18(4):424-432.
4. van der Elst W, van Boxtel MP, van Breukelen GJ, Jolles J. The letter digit substitution test: Normative data for 1,858 healthy participants aged 24-81 from the maastricht aging study (maas): Influence of age, education, and sex. J Clin Exp Neuropsychol. 2006;28(6):998-1009.
5. Lam M, Awasthi S, Watson HJ, Goldstein J, Panagiotaropoulou G, Trubetskoy V et al. Ricopili: Rapid imputation for consortias pipeline. Bioinformatics. 2020;36(3):930-933.
6. Chang CC, Chow CC, Tellier LC, Vattikuti S, Purcell SM, Lee JJ. Second-generation plink: Rising to the challenge of larger and richer datasets. Gigascience. 2015;4:7.
7. Loh PR, Danecek P, Palamara PF, Fuchsberger C, Reshef YA, Finucane HK et al. Reference-based phasing using the haplotype reference consortium panel. Nature Genetics. 2016;48(11):1443-1448.
8. Genomes Project C, Auton A, Brooks LD, Durbin RM, Garrison EP, Kang HM et al. A global reference for human genetic variation. Nature. 2015;526(7571):68-74.
9. Das S, Forer L, Schonherr S, Sidore C, Locke AE, Kwong A et al. Next-generation genotype imputation service and methods. Nat Genet. 2016;48(10):1284-1287.
10. Heger IS, Deckers K, Schram MT, Stehouwer CDA, Dagnelie PC, van der Kallen CJH et al. Associations of the lifestyle for brain health index with structural brain changes and cognition: Results from the maastricht study. Neurology. 2021;97(13):e1300-e1312.
11. Deckers K, van Boxtel MP, Schiepers OJ, de Vugt M, Munoz Sanchez JL, Anstey KJ et al. Target risk factors for dementia prevention: A systematic review and delphi consensus study on the evidence from observational studies. Int J Geriatr Psychiatry. 2015;30(3):234-246.
12. Deckers K, Kohler S, van Boxtel M, Verhey F, Brayne C, Fleming J. Lack of associations between modifiable risk factors and dementia in the very old: Findings from the cambridge city over-75s cohort study. Aging Ment Health. 2018;22(10):1272-1278.
